# Epidemic-Ready Primary Health Care and the Application of the 717 Metrics: A Case Study on a Measles Outbreak in Sierra Leone, January-March 2024

**DOI:** 10.1101/2025.02.24.25322490

**Authors:** Oliver Eleeza, Amy Elizabeth Barrera-Cancedda, Ronald R. Mutebi, Amon Njenga, Mohamed A. Vandi, Stacey Mearns, AbdulRaheem Yakubu, Mame Toure, Susan Michaels-Strasser

## Abstract

The COVID-19 pandemic exposed vulnerabilities in health systems’ abilities to detect, report, and respond to threats. Inadequate preparedness led to healthcare worker infections (HCW), essential service disruptions, and impacts on communities. Primary health care (PHC) is often overlooked in health security initiatives. Epidemic Ready Primary Health Care (ERPHC) is an initiative that strengthens PHC facilities to prevent, detect and respond to outbreaks, while maintaining essential services.

ICAP at Columbia University, the Ministry of Health of Sierra Leone, and Resolve to Save Lives is implementing a multi-year ERPHC project in Sierra Leone. A retrospective data review of 52 confirmed measles cases across four PHC facilities from January – March 2024 was conducted. Data is presented using an adapted 7-1-7 quality improvement approach for detection and notification to evaluate the key tenants of ERPHC: speed, safety, surge.

Out of 52 confirmed cases, 98% and 100% met the first “7” and “1” for detection and notification. Immediate case management and safety actions were completed for all 52 cases (100%). Zero facilities were able to implement two readiness parameters for surge: sufficient supplies and referral pathways. Key bottlenecks included late presentation to health facilities, delayed notification via the electronic case-based surveillance system, inadequate PPE, and no updated referral pathways.

These results underscore the need to scale and implement ERPHC, using adapted 7-1-7 metrics, in PHC facilities. HCW safety, increased community engagement, national supply chain mechanism strengthening, and established patient referral pathways need to be the foci of further health security investment in Sierra Leone.

## Background information

How fast a health system detects and responds to a threat is a critical indicator of its emergency readiness and responsiveness [1, 2]. The COVID-19 pandemic and other recent outbreaks showcased vulnerabilities in health systems’ capacities to be resilient against emerging and re-emerging threats [3]. Healthcare worker (HCW) infections and essential service disruptions are consequences of health system vulnerabilities [4]. In low-resource settings, insufficient preparedness exacerbates existing challenges in fragile health systems, further compromising overall health security [4, 5]. Currently, most health security initiatives focus on national or immediate sub-national health system levels [6]. The role of primary healthcare (PHC) has been mostly overlooked in epidemic planning and preparedness initiatives: a critical oversight given that the entry points for most outbreaks are at the PHC level [6, 7]. Strengthening PHC, specifically the ability to detect, protect against, and respond to threats, can enhance early outbreak detection and response, thereby supporting health system resilience. The 7-1-7 targets were introduced to enable countries to assess and improve outbreak detection and response performance[8]. At the national and sub-national levels of the healthcare system, the approach assesses critical timeliness metrics for pathogens with outbreak potential, emphasizing detection (≤7 days from emergence), notification (target of ≤1 day from detection), and completion of relevant early response actions (target of ≤7 days from notification)[8]. A review of 7-1-7 implementation across five countries identified 61% of the bottlenecks to outbreak detection were at the PHC facility level, with low clinical suspicion among HCWs as the most frequently cited bottleneck[9].

Epidemic Ready Primary Health Care (ERPHC) strengthens PHC systems to effectively prevent, detect and respond to outbreaks, while maintaining essential services. Ensuring facilities can quickly identify cases (speed), manage them safely (safety), and handle increased patient volume (surge). The ERPHC approach builds HCW capacity to connect with communities, rapidly detect and report cases, protect themselves by applying infection prevention control (IPC) measures, and treat cases while maintaining essential services. Key interventions include continuous, real-time mentorship to increase HCW’s clinical index of suspicion for suspected cases of pathogens with outbreak potential, reviewing PHC facility data with HCWs, enhancing HCW IPC knowledge, and ensuring reporting mechanisms for alerts are established. Adapted from the national and sub-national approach, ERPHC uses a modified 7-1-7 tool tailored for the PHC level to assess epidemic readiness and drive improvements. It is applied to all confirmed cases at the facility, providing a broader understanding of performance and enhancing learning opportunities from real-life events. The tool looks at detection and notification timeliness to evaluate the speed of case detection and notification; immediate health facility case management actions to review the safety measures taken for both patients and HCWs; and health facility readiness for further cases to assess preparedness for potential surges in cases.

## Current situation

Sierra Leone has faced many outbreaks, the most notable of these was the Ebola Viral Disease (EVD) outbreak in 2014 [10]. Since this time, there have been considerable investments in the country’s health system architecture to strengthen outbreak preparedness and response. The establishment of an electronic integrated disease surveillance and reporting system (IDSR) and a national IPC program are examples of health system improvements achieved since the end of the EVD outbreak [11–13]. Yet, despite these gains, significant challenges remain. In November 2019, an outbreak of Lassa Fever resulted in the infection of five HCWs [14]. While an epidemiologic link was established among all cases, it took 20 days to detect the index case [15]. After action reviews (AARs) identified gaps including delays in accessing local emergency funding, reporting delays via the electronic case-based surveillance (eCBDS) system, and electronic IDSR (eIDSR) systems, and late distribution of laboratory supplies and PPE [16].

Since January 2024, ICAP at Columbia University (ICAP), in partnership with Resolve to Save Lives (RTSL), the Ministry of Health (MOH), District Health Management Teams (DHMTs), and the National Public Health Agency has implemented the ERPHC model. This case study aims to provide a detailed analysis of the ERPHC model’s implementation in Sierra Leone’s PHC facilities during a measles outbreak utilizing an adapted version of the 7-1-7 metrics. By examining relevant findings and lessons learned, the study seeks to assess the effectiveness of the ERPHC model in enhancing outbreak preparedness and response within PHC facilities.

## Methods

### Design

The study used a retrospective design to assess a measles outbreak that occurred between January 1 and March 31, 2024. An adapted, standardized data collection tool based on the 7-1-7 metrics, incorporating both survey-based and open-ended questions was used to gather information from multiple sources, including IDSR case-based notification forms, the eCBDS system, PHC facility reports, and AAR discussions with HCWs who managed the measles cases. This design allowed for a comprehensive evaluation of outbreak management practices and timelines.

### Setting

The ERPHC model is being implemented across 80 PHC facilities in five districts: Western Area Urban, Western Area Rural, Bombali, Port Loko, and Kenema. These districts collectively represent approximately 43% of Sierra Leone’s total population [17].

This case study examines the implementation of the ERPHC model in two of the ERPHC supported districts— Port Loko and Bombali —focusing on four PHC facilities that managed confirmed measles cases. Amongst these PHC facilities, there were 2 Community Health Centers (CHCs), which typically have 10-15 staff members and offer preventive and curative services with a population of 10,000 to 30,000 or more within 15 km, 2 Community Health Posts (CHPs), which offer basic maternal and child health services to 5,000 to 10,000 individuals within 8 km (5 miles) radius and has 1-3 staff members[18].

In Sierra Leone, PHC facilities utilize multiple channels to report suspected measles cases and other priority pathogens identified by HCWs. Alerts can be communicated via text message (SMS), WhatsApp, or phone calls to higher levels of the health system, such as the DHMT and the central MOH. Additionally, all suspected cases of priority pathogens can be directly entered into the eCBDS or weekly aggregate eIDSR systems.

### Population

A total of 52 measles cases detected at four PHCs comprised the population. This case study focused on cases that met the following criteria:

Inclusion Criteria: Suspected measles cases that were subsequently confirmed and detected at PHC facility implementing the ERPHC model from January 1 – March 31, 2023.

Exclusion Criteria: Cases were excluded if they did not receive confirmation of measles or were outside the ERPHC facility catchment area.

### Data collection Tool

A standardized data collection tool, adapted from the 7-1-7 outbreak assessment framework, was used in this case study to assess PHC facility readiness (see Annex 1). The tool was organized into four key sections: 1) background information; 2) timeliness metrics for detection and notification; 3) immediate case management and safety actions; and 4) facility readiness to manage a surge in cases, assessed through ten critical readiness actions. By identifying strengths, gaps, and areas for improvement, the tool provided actionable insights to enhance PHC preparedness and response capabilities.

### Data collection Process

Data collection was conducted by a team of two ICAP staff, two DHMT IPC focal persons, and two IDSR focal persons. They used the data collection tool to extract information on measles cases from PHC facility registers, case-based notification forms, and the eCBDS system. Additionally, nine HCWs, including four facility managers and five nurses who managed suspected cases, were interviewed during an AAR using survey-based and open-ended questions. These focused on safety measures and surge capacity. Data were collected over five days at four PHC facilities, initially recorded on paper, and later entered into Microsoft Excel for analysis. Prior to data collection, ICAP and DHMT staff participated in a one-day orientation to ensure consistency in tool use, data abstraction, and interview techniques.

### Data management

The dataset was reviewed to ensure completeness, accuracy, and logical consistency. At the PHC level, all required fields were populated according to predefined specifications, with no critical information omitted. The data collection team verified PHC facility records with the eCBDS system to minimize discrepancies. Transcription errors were identified and corrected, while analytical checks flagged and resolved inconsistencies and anomalies. No personally identifiable information was collected, and all data were anonymized, securely stored, and accessible only to authorized personnel.

### Analysis

Descriptive statistics were used to calculate the number of cases meeting the targets for timely detection (Dd-Do ≤7 days) and notification (Dn – Dd ≤1 day) (see Box 1).

#### Box 1: Definitions for measures of detection and notification (Speed)

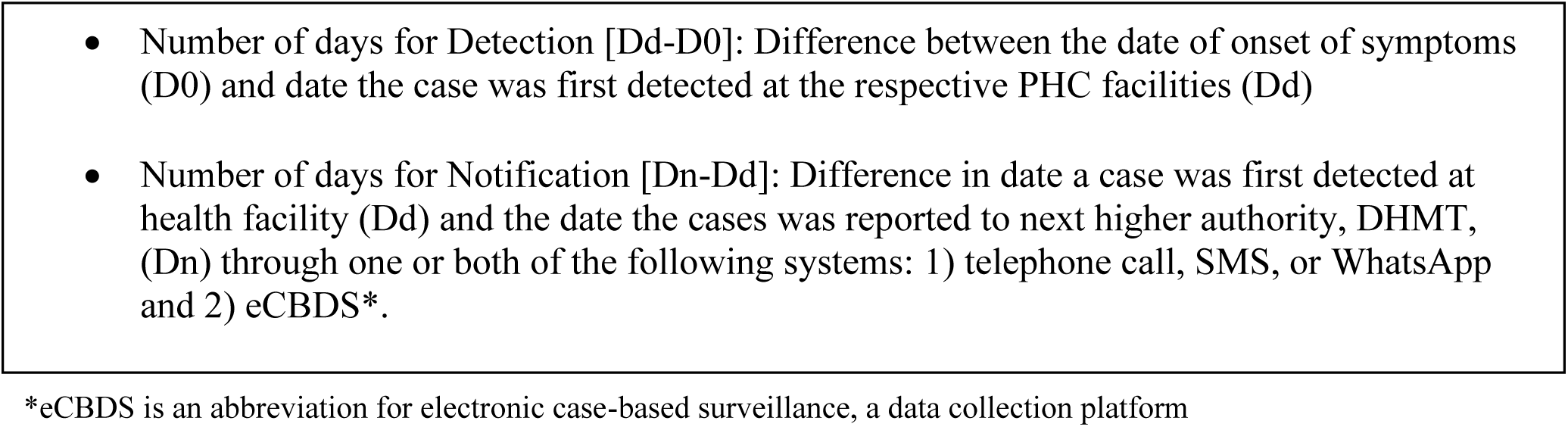

To assess safety, descriptive statistics were used to calculate the number of cases that: 1.) were isolated at the PHC, 2.) had HCWs use the appropriate PPE for a suspected case of measles, and 3.) received correct actions for clinical case management (see Annex 1). Additionally, HCWs’ perceptions of safety requirements were analyzed using thematic analysis.

To assess surge readiness capacity, calculations on the proportion of PHC facilities that met each of the 10 requirements for PHC surge readiness during the measles outbreak were conducted (see Annex 1). Additionally, HCWs’ perceptions on surge requirements were analyzed using thematic analysis.

All data collected using the data collection tool for each PHC facility can be reviewed in Annexes 2-4.

## Results

A total of 52 measles cases were detected in PHC facilities implementing ERPHC. Of the 80 PHCs implementing the ERPHC model, four PHC facilities received measles cases. One PHC facility detected 49 confirmed cases representing 94% of all 52 cases assessed, with an average volume of two cases per day [median: 1 (1-5)]. The remaining three PHC facilities had one confirmed case each (6%). Table 1 below provides details of PHC facilities performance on detection and notification metrics (speed).

**Table 1:**
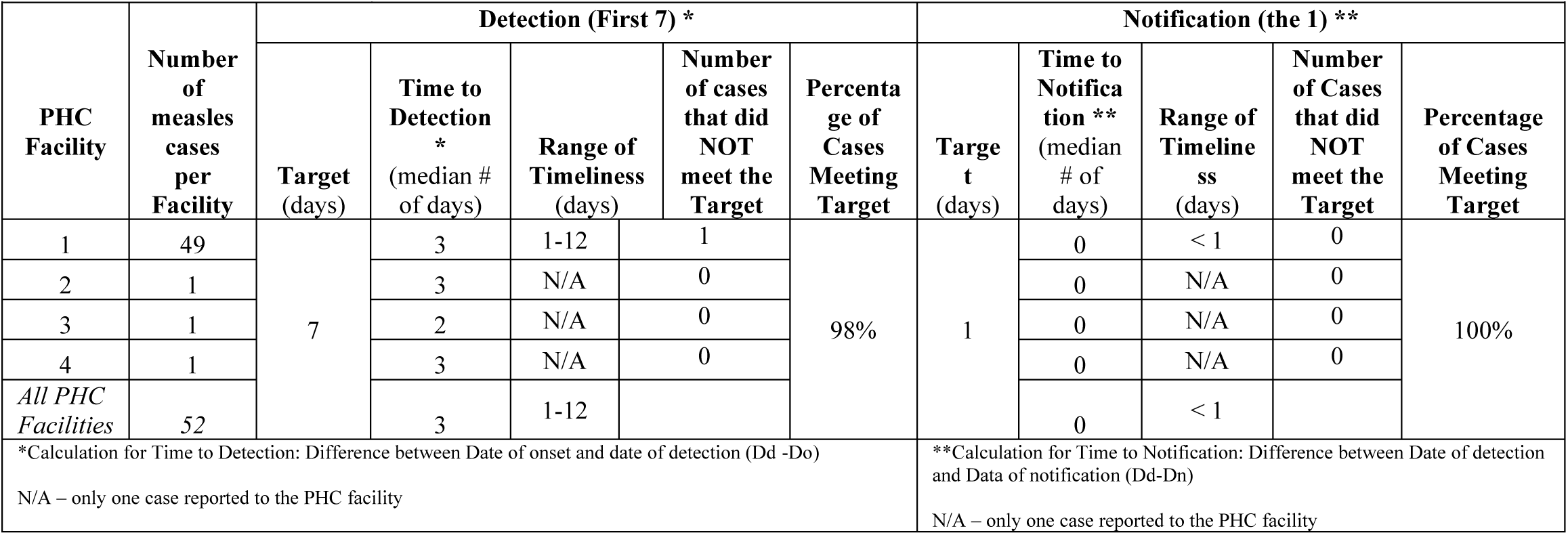
Analysis of Metrics for Measles Outbreak in Four PHC Facilities in Sierra Leone.

Of the 52 confirmed cases, 51 (98%) met the target for the “first 7” of the 7-1-7 metrics for detection. (see Table 1). The PHC with the most cases (n=49) had a range for detection of 1-12 days. This PHC reported a single case on day 12, when the suspected case presented to the PHC and had already had symptoms for 12 days prior. The other 48 cases at this PHC facility were detected within the 7-day range, making the median time to detection for all four PHC facilities 3 days.

Of the 52 confirmed measles cases, 52 (100%) met the “the 1” of the 7-1-7 metrics for notification. All alerts were notified via SMS, WhatsApp message, or phone call to the DHMT. All alerts were eventually notified via the eCBDS system, though not within the targeted one-day timeframe. Only 44% (23/52) met “the 1” target using the eCBDS system, which directly notifies the national level of a threat. Reasons for these delays included HCW workload, technological challenges and limited connectivity.

Table 2: depicts the number of cases (n=52) where immediate case management actions to ensure HCW and patient safety were implemented. Specifically, HCWs placed measles cases into isolation (n=52, 100%)) donned the correct PPE (n=52, 100%), and provided initial treatment based on SOPs (n=52, 100%) (Table 2). The IPC job aides, SOPs, and HCWs’ training, and case management protocols were identified as enablers to successful implementation of case management safety actions.

**Table 2:**
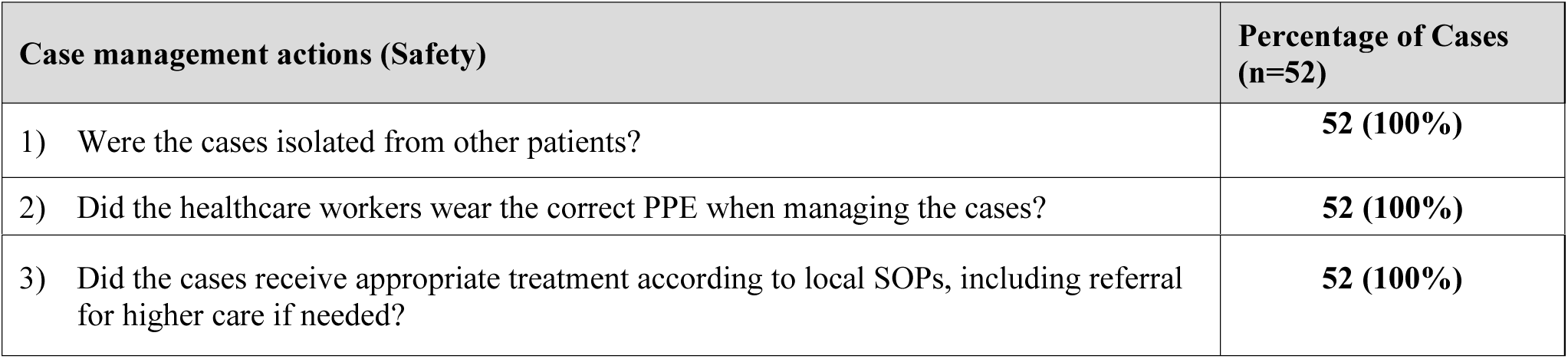
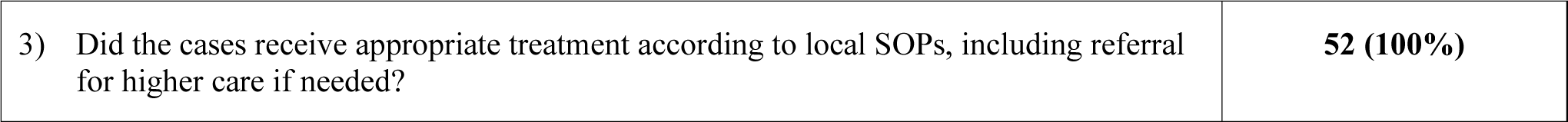
Number of Cases where Immediate Case Management Actions for HCW and Patient Safety were Implemented in Four PHCs in Sierra Leone.

| Case management actions (Safety) | Percentage of Cases (n=52) |
| --- | --- |
| 1) Were the cases isolated from other patients? | 52 (100%) |
| 2) Did the healthcare workers wear the correct PPE when managing the cases? | 52 (100%) |
| 3) Did the cases receive appropriate treatment according to local SOPs, including referral for higher care if needed? | **52 (100%)** |

Table 3 details the surge parameters for HF readiness at the four PHC facilities that detected measles cases.

**Table 3:**
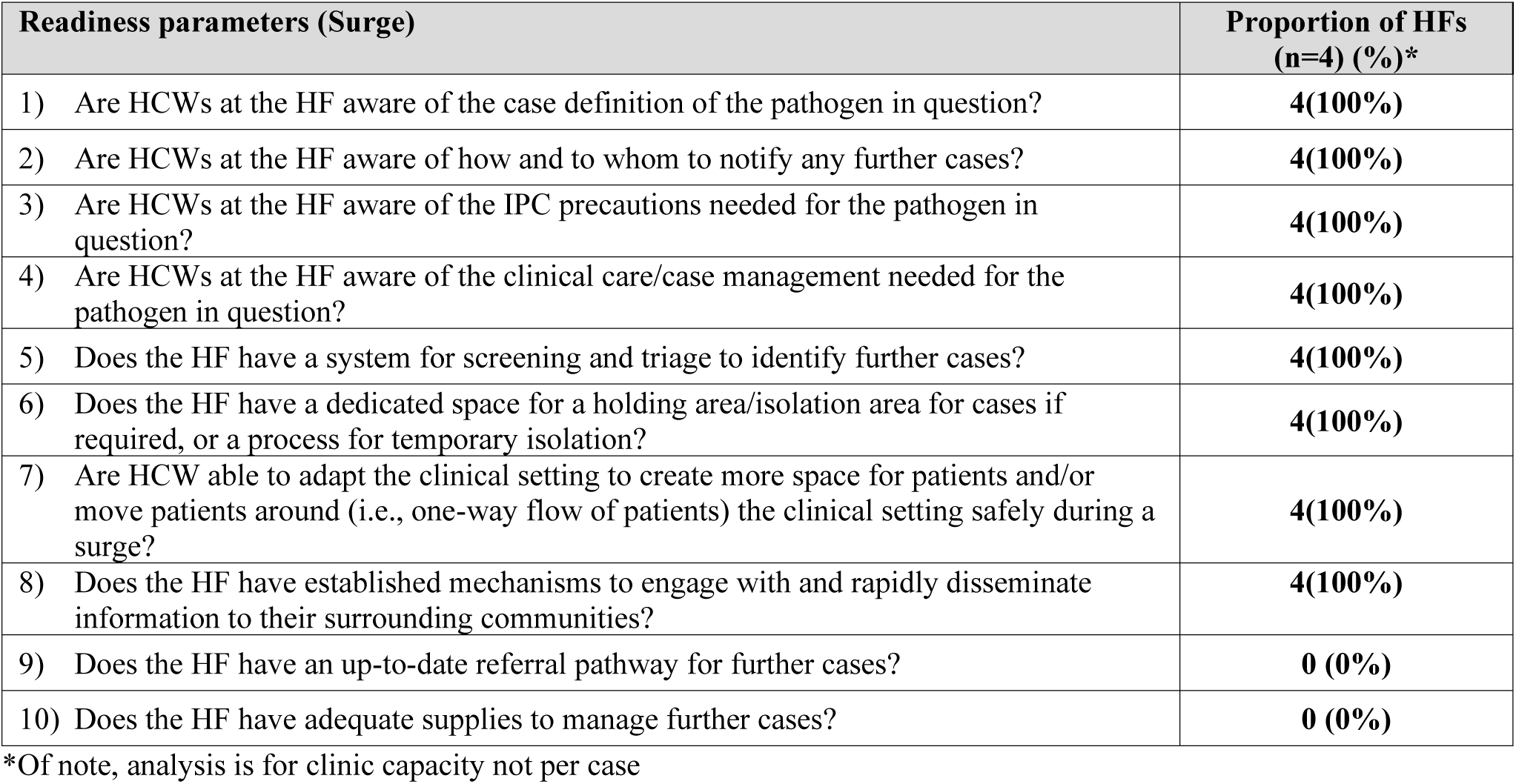
Proportion of PHCs meeting the immediate case management actions (safety) and readiness parameters (surge) for a measle outbreak in four PHCs implementing the ERPHC model.

All four (100%) PHC facilities were able to implement readiness parameters for surge requirements 1-8. None of the PHC facilities were able to implement readiness parameters for surge requirements 9 and 10. Supply chain issues, limited PPE availability, and no referral pathways were identified as key bottlenecks.

In discussion with HCWs during an AAR, HCWs stated that not having PPE supplies was the top bottleneck for keeping them safe. HCWs reported that not having adequate supplies, including PPE, as well as a lack of updated referral pathways at the PHC were key bottlenecks for surge capacity. HCWs identified IPC mentorship that allowed for an increase in clinical suspicion of a priority pathogen, SOPs/job aides, and case management protocols as implementation enablers.

## Discussion

Primary healthcare facilities, implementing the ERPHC model, were largely successful in achieving the detection and notification targets of the 7-1-7 metrics adapted for PHCs. This achievement shows that with increased investment in continuous, real-time mentorship at the PHC level, HCWs can improve their clinical index of suspicion and rapidly identify and notify a suspected case of a pathogen of outbreak potential.

Regarding speed of detection (“first 7”), the health facility reporting the highest number of cases (49) had a detection range of 1–12 days. Notably, only one patient presented on day 12, reflecting a delay in seeking care following symptom onset. Despite this delay, HCWs at the facility promptly suspected measles upon the patient’s arrival and notified the case. The remaining 48 cases were detected within the critical window of 1–7 days, resulting in a median detection time of 3 days for the facility. All confirmed measles cases were successfully reported within 1 day using SMS, WhatsApp, or phone. However, only 44% percent of the alerts were notified within 1 day via the eCBDS or eIDSR platforms, which go directly to the national level - that can initiate rapid response teams (RRT), emergency funds, and declare an outbreak – for *response* initiation. Reasons for this lack of eCBDS utilization by HCWs was related to technological challenges, insufficient internet capacity, and the three other options (SMS, WhatsApp, phone) allow for reporting mechanisms to be returned to the HCW. Messages of receipt and dialogue with another individual at the DHMT may provide reassurance and encouragement to HCWs, compared to entering information into the electronic platforms, where the imported information is sent off and no information returned to the HCW. This lack of returned communication to HCWs using the eCBDS system, while reporting on the same pathogen over and again during an outbreak, may lead to report fatigue. Human-to-human communication may be the key to increase HCW reporting.

The safety actions were also achieved with great success in this case study, mostly likely attributed to the integrated interventions of the ERPHC model. The ERPHC approach integrates surveillance, IPC, and case management into mentoring to ensure that HCWs are equipped to detect and report cases, understand the necessary IPC measures to ensure their safety, and provide appropriate treatment to patients. These three areas: surveillance, IPC, and case management, are typically siloed and focused on independently of the other. Based on the results of this case study, integrating them together better equips HCWs at PHCs for outbreaks.

Regarding the two surge actions that no PHC facility successfully implemented—adequate supplies and a referral pathway for cases—these shortcomings may be linked to challenges in national supply chain mechanisms and the absence of SOPs during the early months of ERPHC implementation. During outbreaks, HCWs often lack sufficient PPE to ensure their own safety and that of their patients. While PHC facilities implementing the ERPHC model are not immune to these challenges, this should not be accepted as the norm.

### Limitations

There were limitations to this case study. First, retrospective data collection may have led to recall bias by HCWs at the PHC facilities during the AAR. Second, the PHCs in this case study may not be reflective of PHCs in Sierra Leone, in that they received interventions that are not routine in other PHC facilities. Thus, these results may not be generalizable to other PHC facilities. Despite these limitations, the insight this case study provides can better prepare PHC facilities for outbreaks

### Recommendations and Conclusions

This case study demonstrates the effective application of the adapted 7-1-7 metrics at the PHC level, showing that these metrics, traditionally employed at national and subnational levels, can be successfully modified and implemented at lower tiers of the health system to assess and enhance epidemic readiness. To strengthen preparedness for future outbreaks, the ERPHC model should be prioritized, scaled up, and adequately resourced as a fundamental health security intervention in Sierra Leone. Expanding this approach at the PHC level is essential for building resilient health systems and ensuring timely, effective outbreak responses.

Community engagement is a cornerstone of effective outbreak response. Delayed presentation to PHC facilities exacerbates transmission, highlighting the importance of ensuring that communities are promptly informed of outbreaks. Establishing direct links between communities and PHCs—through community health workers or focal points—enables the rapid dissemination of risk communication and educational materials at the onset of an outbreak. Reducing the time between symptom onset, case detection, and notification of epidemic-prone pathogens is crucial for expediting response actions, enhancing outbreak control efforts, and preventing further transmission.

Ensuring HCW safety through comprehensive interventions is also critical. This case study underscores the need to proactively address PHC infrastructural gaps and supply chain challenges ahead of outbreaks to mitigate their impact. PHCs must have reliable communication systems, internet connectivity, and functional technologies for the swift reporting of suspected cases. Beyond infrastructure, HCWs must feel supported by these systems, receiving timely, reliable guidance to manage suspected cases effectively, particularly under stressful conditions. To prevent overcrowding and minimize HCW exposure to high-risk environments, robust referral pathways should be established to appropriately direct patient care. Additionally, addressing supply chain issues is essential, as limited access to personal protective equipment (PPE) places HCWs at increased risk of infection. Prepositioning PPE at PHC facilities ensures that HCWs can protect themselves when managing suspected cases of epidemic-prone pathogens.

Prioritizing community engagement and HCW safety measures is vital for achieving strong outbreak preparedness and enhancing health security. The early results from the ERPHC model in Sierra Leone demonstrate its potential to drive significant improvements in preparedness actions at the PHC level. This case study illustrates the benefits of adapting and utilizing the 7-1-7 metrics at health facilities to better understand performance, readiness, and to drive improvements. By fostering timely, proactive interventions, the ERPHC model can further bolster health system resilience, address systemic vulnerabilities, and improve the capacity to manage outbreaks effectively. Ultimately, these early actions are crucial to ensuring that Sierra Leone protects its most valuable asset during outbreaks: its people.

## Annexes

**Annex 1:**
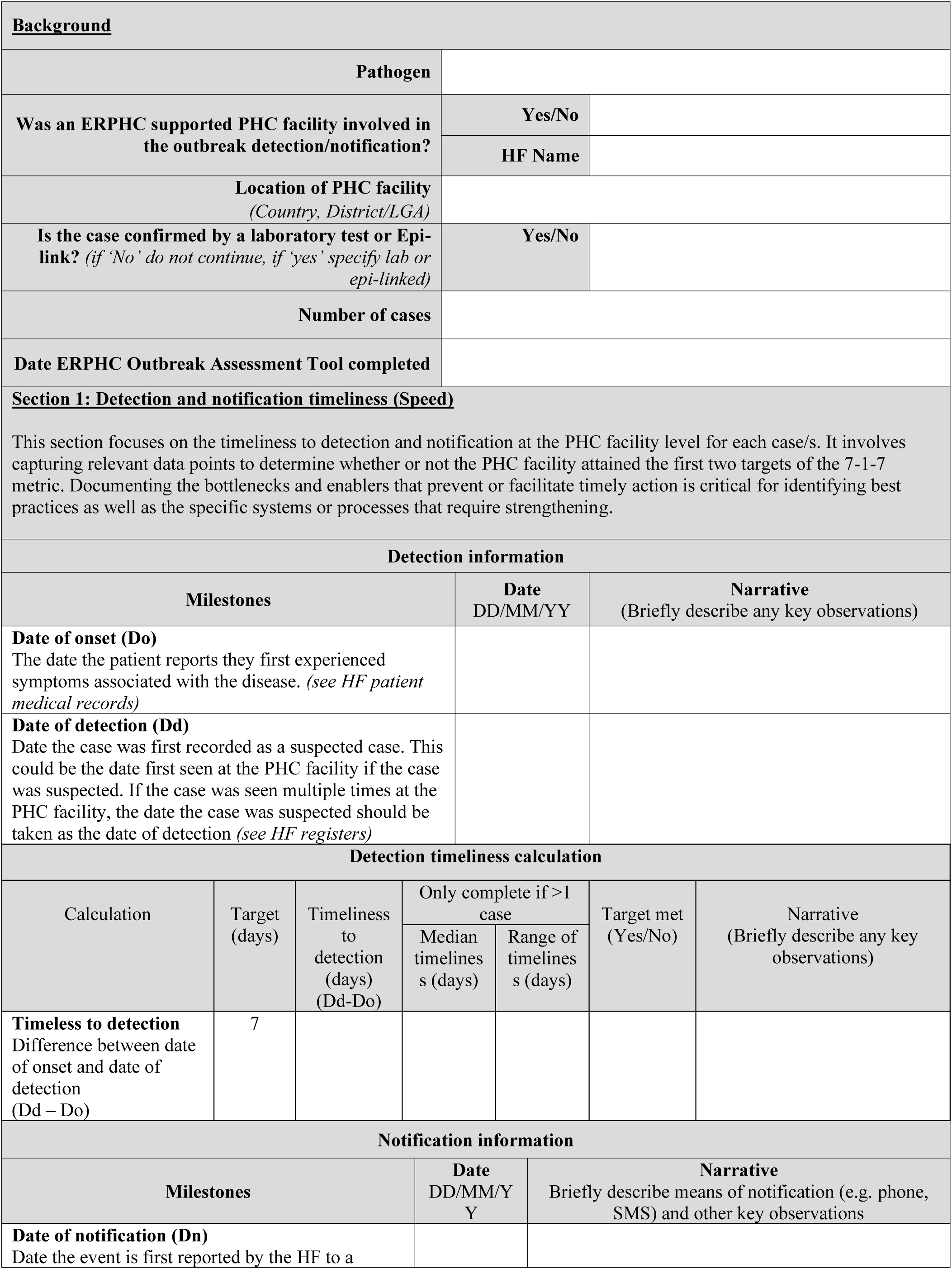
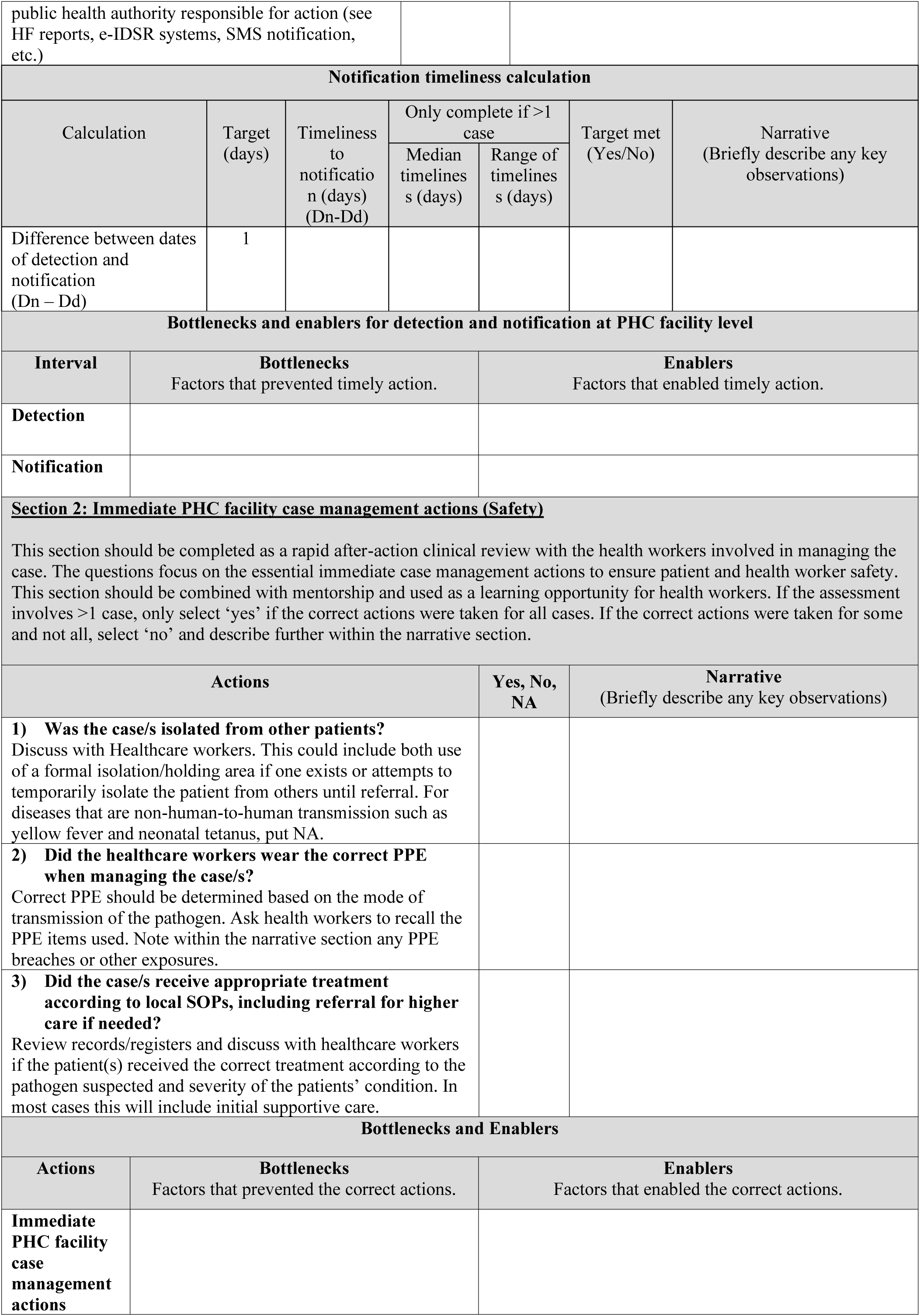
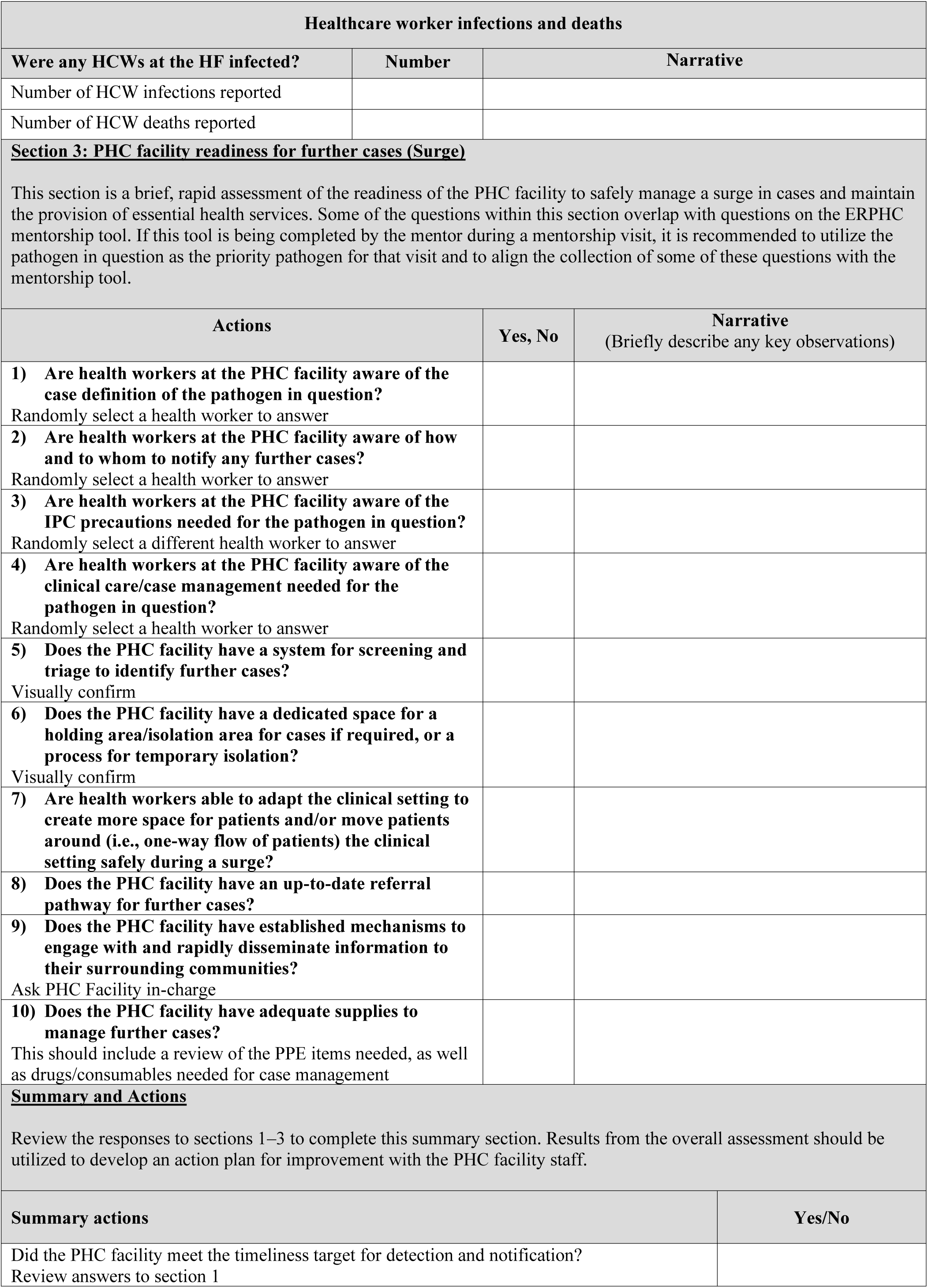
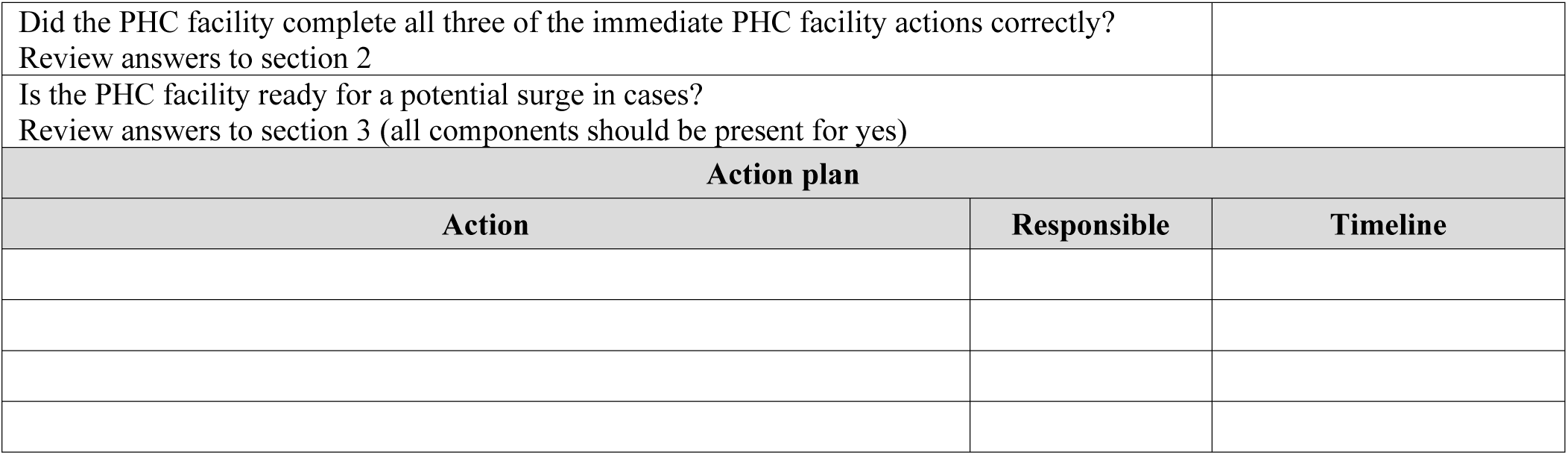
ERPHC Outbreak Assessment Tool.

| Background |  |  |  |  |  |  |
| --- | --- | --- | --- | --- | --- | --- |
| Pathogen |  |  |  |  |  |  |
| Was an ERPHC supported PHC facility involved in the outbreak detection/notification? |  |  | Yes/No |  |  |  |
|  |  |  | HF Name |  |  |  |
| Location of PHC facility<br>(Country, District/LGA) |  |  |  |  |  |  |
| Is the case confirmed by a laboratory test or Epi-link? (if 'No' do not continue, if 'yes' specify lab or epi-linked) |  |  | Yes/No |  |  |  |
| Number of cases |  |  |  |  |  |  |
| Date ERPHC Outbreak Assessment Tool completed |  |  |  |  |  |  |
| Section 1: Detection and notification timeliness (Speed) |  |  |  |  |  |  |
| This section focuses on the timeliness to detection and notification at the PHC facility level for each case/s. It involves capturing relevant data points to determine whether or not the PHC facility attained the first two targets of the 7-1-7 metric. Documenting the bottlenecks and enablers that prevent or facilitate timely action is critical for identifying best practices as well as the specific systems or processes that require strengthening. |  |  |  |  |  |  |
| Detection information |  |  |  |  |  |  |
| Milestones |  |  | Date<br>DD/MM/YY | Narrative<br>(Briefly describe any key observations) |  |  |
| **Date of onset (Do)**<br>The date the patient reports they first experienced symptoms associated with the disease. (see HF patient medical records) |  |  |  |  |  |  |
| **Date of detection (Dd)**<br>Date the case was first recorded as a suspected case. This could be the date first seen at the PHC facility if the case was suspected. If the case was seen multiple times at the PHC facility, the date the case was suspected should be taken as the date of detection (see HF registers) |  |  |  |  |  |  |
| Detection timeliness calculation |  |  |  |  |  |  |
| Calculation | Target<br>(days) | Timeliness<br>to<br>detection<br>(days)<br>(Dd-Do) | Only complete if >1<br>case |  | Target met<br>(Yes/No) | Narrative<br>(Briefly describe any key<br>observations) |
|  |  |  | Median<br>timelines<br>s (days) | Range of<br>timelines<br>s (days) |  |  |
| **Timeless to detection**<br>Difference between date<br>of onset and date of<br>detection<br>(Dd – Do) | 7 |  |  |  |  |  |
| Notification information |  |  |  |  |  |  |
| Milestones |  |  | Date<br>DD/MM/Y<br>Y | Narrative<br>Briefly describe means of notification (e.g. phone, SMS) and other key observations |  |  |
| **Date of notification (Dn)**<br>Date the event is first reported by the HF to a |  |  |  |  |  |  |

| public health authority responsible for action (see HF reports, e-IDSR systems, SMS notification, etc.) |  |  |  |  |  |  |
| --- | --- | --- | --- | --- | --- | --- |
| **Notification timeliness calculation** |  |  |  |  |  |  |
| Calculation | Target (days) | Timeliness to notification (days) (Dn-Dd) | Only complete if >1 case |  | Target met (Yes/No) | Narrative (Briefly describe any key observations) |
|  |  |  | Median timelines (days) | Range of timelines (days) |  |  |
| Difference between dates of detection and notification (Dn – Dd) | 1 |  |  |  |  |  |
| **Bottlenecks and enablers for detection and notification at PHC facility level** |  |  |  |  |  |  |
| Interval | Bottlenecks<br>Factors that prevented timely action. |  |  | Enablers<br>Factors that enabled timely action. |  |  |
| **Detection** |  |  |  |  |  |  |
| **Notification** |  |  |  |  |  |  |
| **<u>Section 2: Immediate PHC facility case management actions (Safety)</u>** |  |  |  |  |  |  |
| This section should be completed as a rapid after-action clinical review with the health workers involved in managing the case. The questions focus on the essential immediate case management actions to ensure patient and health worker safety. This section should be combined with mentorship and used as a learning opportunity for health workers. If the assessment involves >1 case, only select 'yes' if the correct actions were taken for all cases. If the correct actions were taken for some and not all, select 'no' and describe further within the narrative section. |  |  |  |  |  |  |
| Actions |  |  | Yes, No, NA | Narrative<br>(Briefly describe any key observations) |  |  |
| **1) Was the case/s isolated from other patients?**<br>Discuss with Healthcare workers. This could include both use of a formal isolation/holding area if one exists or attempts to temporarily isolate the patient from others until referral. For diseases that are non-human-to-human transmission such as yellow fever and neonatal tetanus, put NA. |  |  |  |  |  |  |
| **2) Did the healthcare workers wear the correct PPE when managing the case/s?**<br>Correct PPE should be determined based on the mode of transmission of the pathogen. Ask health workers to recall the PPE items used. Note within the narrative section any PPE breaches or other exposures. |  |  |  |  |  |  |
| **3) Did the case/s receive appropriate treatment according to local SOPs, including referral for higher care if needed?**<br>Review records/registers and discuss with healthcare workers if the patient(s) received the correct treatment according to the pathogen suspected and severity of the patients' condition. In most cases this will include initial supportive care. |  |  |  |  |  |  |
| **Bottlenecks and Enablers** |  |  |  |  |  |  |
| Actions | Bottlenecks<br>Factors that prevented the correct actions. |  |  | Enablers<br>Factors that enabled the correct actions. |  |  |
| **Immediate PHC facility case management actions** |  |  |  |  |  |  |

| Healthcare worker infections and deaths |  |  |
| --- | --- | --- |
| Were any HCWs at the HF infected? | Number | Narrative |
| Number of HCW infections reported |  |  |
| Number of HCW deaths reported |  |  |
| **Section 3: PHC facility readiness for further cases (Surge)**<br>This section is a brief, rapid assessment of the readiness of the PHC facility to safely manage a surge in cases and maintain the provision of essential health services. Some of the questions within this section overlap with questions on the ERPHC mentorship tool. If this tool is being completed by the mentor during a mentorship visit, it is recommended to utilize the pathogen in question as the priority pathogen for that visit and to align the collection of some of these questions with the mentorship tool. |  |  |
| Actions | Yes, No | Narrative<br>(Briefly describe any key observations) |
| **1) Are health workers at the PHC facility aware of the case definition of the pathogen in question?**<br>Randomly select a health worker to answer |  |  |
| **2) Are health workers at the PHC facility aware of how and to whom to notify any further cases?**<br>Randomly select a health worker to answer |  |  |
| **3) Are health workers at the PHC facility aware of the IPC precautions needed for the pathogen in question?**<br>Randomly select a different health worker to answer |  |  |
| **4) Are health workers at the PHC facility aware of the clinical care/case management needed for the pathogen in question?**<br>Randomly select a health worker to answer |  |  |
| **5) Does the PHC facility have a system for screening and triage to identify further cases?**<br>Visually confirm |  |  |
| **6) Does the PHC facility have a dedicated space for a holding area/isolation area for cases if required, or a process for temporary isolation?**<br>Visually confirm |  |  |
| **7) Are health workers able to adapt the clinical setting to create more space for patients and/or move patients around (i.e., one-way flow of patients) the clinical setting safely during a surge?** |  |  |
| **8) Does the PHC facility have an up-to-date referral pathway for further cases?** |  |  |
| **9) Does the PHC facility have established mechanisms to engage with and rapidly disseminate information to their surrounding communities?**<br>Ask PHC Facility in-charge |  |  |
| **10) Does the PHC facility have adequate supplies to manage further cases?**<br>This should include a review of the PPE items needed, as well as drugs/consumables needed for case management |  |  |
| **Summary and Actions**<br>Review the responses to sections 1–3 to complete this summary section. Results from the overall assessment should be utilized to develop an action plan for improvement with the PHC facility staff. |  |  |
| **Summary actions** |  | **Yes/No** |
| Did the PHC facility meet the timeliness target for detection and notification?<br>Review answers to section 1 |  |  |

|  |  |
| --- | --- |
| Did the PHC facility complete all three of the immediate PHC facility actions correctly?<br>Review answers to section 2 |  |
| Is the PHC facility ready for a potential surge in cases?<br>Review answers to section 3 (all components should be present for yes) |  |
| **Action plan** |  |
| **Action** | **Responsible** |

**Annex 2:** PHC 1 Measles Outbreak 7-1-7 Analysis Results.

| **<u>Background</u>** |  |  |
| --- | --- | --- |
| **Pathogen** | **Measles** |  |
| **Was an ERPHC supported primary healthcare (PHC) facility involved in the outbreak detection/notification?** | **Yes/No** |  |
|  | **HF Name** | **PHC 1** |
| **Location of PHC facility**<br>(Country, District/LGA) |  |  |
| **Is the case confirmed by a laboratory test or Epi-link?** (if 'No' do not continue, if 'yes' specify lab or epi-linked) | **Yes/No** | **Yes** |
| **Number of cases** | **49** |  |
| **Date ERPHC Outbreak Assessment Tool completed** |  |  |
| **<u>Section 1: Detection and notification timeliness (Speed)</u>** |  |  |
| This section focuses on the timeliness to detection and notification at the PHC facility level for each case/s. It involves capturing relevant data points to determine whether or not the PHC facility attained the first two targets of the 7-1-7 metric. Documenting the bottlenecks and enablers that prevent or facilitate timely action is critical for identifying best practices as well as the specific systems or processes that require strengthening. |  |  |
| **Detection information** |  |  |
| **Milestones** | **Date**<br>DD/MM/YY | **Narrative**<br>(Briefly describe any key observations) |
| **Date of onset (Do)**<br>The date the patient reports they first experienced symptoms associated with the disease. (see HF patient medical records) |  | The cases cover a range of dates of onset from December 28, 2023, to March 16, 2024, the majority of the cases fall within the months of January and February 2024, suggesting a potential clustering or higher incidence during these two months. |
| **Date of detection (Dd)**<br>Date the case was first recorded as a suspected case. This could be the date first seen at the PHC facility if the case was suspected. If the case was seen multiple times at the PHC facility, the date the case was suspected should be taken as the date of detection (see HF registers) |  | All 49 cases presented at the HF were recorded in the HF registers and in the case-base notification forms, where all the cases dates of detection/ seen at HF are recorded. |
| **Detection timeliness calculation** |  |  |

| Calculation | Target (days) | Timeliness to detection (days) (Dd-Do) | Only complete if >1 case |  | Target met (Yes/No) | Narrative (Briefly describe any key observations) |
| --- | --- | --- | --- | --- | --- | --- |
|  |  |  | Median timeliness (days) | Range of timeliness (days) |  |  |
| **Timeless to detection**<br>Difference between date of onset and date of detection (Dd – Do) | 7 | Case 1: 3 days<br>Case 2: 7 days<br>Case 3: 2 days<br>Case 4: 3 days<br>Case 5: 4 days<br>Case 6: 2 days<br>Case 7: 5 days<br>Case 8: 2 days<br>Case 9: 2 days<br>Case 10: 1 day<br>Case 11: 2 days<br>Case 12: 3 days<br>Case 13: 2 days<br>Case 14: 7 days<br>Case 15: 4 days<br>Case 16: 1 days<br>Case 17: 2 days<br>Case 18: 3 days<br>Case 19: 2 days<br>Case 20: 3 days<br>Case 21: 2 days<br>Case 22: 3 days<br>Case 23: 2 days<br>Case 24: 3 days<br>Case 25: 3 days<br>Case 26: 3 days<br>Case 27: 2 days<br>Case 28: 1 day<br>Case 29: 2 days<br>Case 30: 3 days<br>Case 31: 3 days<br>Case 32: 3 days<br>Case 33: 2 days<br>Case 34: 3 days<br>Case 35: 2 days<br>Case 36: 4 days<br>Case 37: 2 days<br>Case 38: 3 days<br>Case 39: 2 days<br>Case 40: 2 days<br>Case 41: 2 days<br>Case 42: 4 days<br>Case 43: 6 days<br>Case 44: 3 days<br>Case 45: 5 days<br>Case 46: 12 days<br>Case 47: 3 days<br>Case 48: 4 days<br>Case 49: 2 days | 3 | 1-12 | Yes | 1/49 cases presented to the HF after 12 days of onset. The remaining 48/49 cases presented to the HF between 1-7 days for care. |
| Notification information |  |  |  |  |  |  |
| Milestones |  | Date DD/MM/YY |  | Narrative<br>Briefly describe means of notification (e.g. phone, SMS) and other key observations |  |  |
| **Date of notification (Dn)**<br>Date the event is first reported by the HF to a public health authority |  |  |  | All 49 cases were notified immediately by a phone call, SMS, or WhatsApp message to the District Surveillance Officers (DSO). Of the 49 cases, 23 |  |  |

| responsible for action (see HF reports, e-IDSr systems, SMS notification, etc.) |  | cases were also immediately entered into the eCBDS. The remaining 26/49 cases were entered into the eCBDS after 14-83 days resulting in a long notification period via the eCBDS system. |  |  |  |  |
| --- | --- | --- | --- | --- | --- | --- |
| Notification timeliness calculation |  |  |  |  |  |  |
| Calculation | Target (days) | Timeliness to notification (days) (Dn-Dd) | Only complete if >1 case |  | Target met (Yes/No) | Narrative (Briefly describe any key observations) |
|  |  |  | Median timeliness (days) | Range of timeliness (days) |  |  |
| Difference between dates of detection and notification (Dn – Dd) | 1 | Case 1: 0 day<br>Case 2:0 day<br>Case 3: 0 day<br>Case 4: 0 day<br>Case 5: 0 day<br>Case 6: 0 day<br>Case 7: 0 day<br>Case 8: 0 day<br>Case 9: 0 day<br>Case 10: 0 day<br>Case 11: 0 day<br>Case 12: 0 day<br>Case 13: 0 day<br>Case 14: 0 day<br>Case 15: 0 day<br>Case 16: 0 day<br>Case 17: 0 day<br>Case 18: 0 day<br>Case 19: 0 day<br>Case 20: 0 day<br>Case 21: 0 day<br>Case 22: 0 day<br>Case 23 : 0 day<br>Case 24: 0 day<br>Case 25: 0 day<br>Case 26: 0 day<br>Case 27: 0 day<br>Case 28: 0 day<br>Case 29: 0 day<br>Case 30: 0 day<br>Case 31: 0 day<br>Case 32: 0 day<br>Case 33: 0 day<br>Case 34: 0 day<br>Case 35: 0 day<br>Case 36: 0 day<br>Case 37: 0 day<br>Case 38: 0 day<br>Case 39: 0 day<br>Case 40: 0 day<br>Case 41: 0 day<br>Case 42: 0 day<br>Case 43: 0 day<br>Case 44: 0 day<br>Case 45: 0 day<br>Case 46: 0 day<br>Case 47:0 day<br>Case 48:0 day<br>Case 49:0 day | 0 | <1 day | Yes | All cases were reported within a day of detection. |
| Bottlenecks and enablers for detection and notification at PHC facility level |  |  |  |  |  |  |

| Interval | Bottlenecks<br>Factors that prevented timely action. | Enablers<br>Factors that enabled timely action. |  |
| --- | --- | --- | --- |
| Detection | Late presentation of some cases to the HFs | Health workers knowledge of case definition |  |
| Notification | Some cases not immediately reported into eCBDS system due to knowledge gap, unclear roles, tablet, or connectivity issues | Availability and knowledge of alternative means of notification e.g.) phone call, SMS |  |
| **<u>Section 2: Immediate PHC facility case management actions (Safety)</u>**<br>This section should be completed as a rapid after-action clinical review with the health workers involved in managing the case. The questions focus on the essential immediate case management actions to ensure patient and health worker safety. This section should be combined with mentorship and used as a learning opportunity for health workers. If the assessment involves >1 case, only select 'yes' if the correct actions were taken for all cases. If the correct actions were taken for some and not all, select 'no' and describe further within the narrative section. |  |  |  |
| Actions |  | Yes, No, NA | Narrative<br>(Briefly describe any key observations) |
| **1) Was the case/s isolated from other patients?**<br>Discuss with Healthcare workers. This could include both use of a formal isolation/holding area if one exists or attempts to temporarily isolate the patient from others until referral. For diseases that are non-human-to-human transmission such as yellow fever and neonatal tetanus, put NA. |  | Yes | HF isolated the cases in their dedicated isolation area (permanent space) as required. . |
| **2) Did the healthcare workers wear the correct PPE when managing the case/s?**<br>Correct PPE should be determined based on the mode of transmission of the pathogen. Ask health workers to recall the PPE items used. Note within the narrative section any PPE breaches or other exposures. |  | Yes | All HFs used appropriate PPEs while managing the cases, although the HF reported inadequate PPEs in case of an acute surge in cases. |
| **3) Did the case/s receive appropriate treatment according to local SOPs, including referral for higher care if needed?**<br>Review records/registers and discuss with healthcare workers if the patient(s) received the correct treatment according to the pathogen suspected and severity of the patients' condition. In most cases this will include initial supportive care. |  | Yes | All 49 cases were treated on the day of visiting the HFs |
| **Bottlenecks and Enablers** |  |  |  |
| Actions | Bottlenecks<br>Factors that prevented the correct actions. | Enablers<br>Factors that enabled the correct actions. |  |
| **Immediate PHC facility case management actions** | Limited availability of PPE supplies<br>availability at HF | Availability of case management SOPs<br>Availability of IPC job aides and SOPs |  |
| **Healthcare worker infections and deaths** |  |  |  |
| Were any HCWs at the HF infected? | Number | Narrative |  |
| Number of HCW infections reported | 0 | No HCW Infected |  |
| Number of HCW deaths reported | 0 | No HCW died |  |
| **<u>Section 3: PHC facility readiness for further cases (Surge)</u>**<br>This section is a brief, rapid assessment of the readiness of the PHC facility to safely manage a surge in cases and maintain the provision of essential health services. Some of the questions within this section overlap with questions on the ERPHC mentorship tool. If this tool is being completed by the mentor during a mentorship visit, it is recommended to utilize the pathogen in question as the priority pathogen for that visit and to align the collection of some of these questions with the mentorship tool. |  |  |  |

| **Actions** | **Yes, No** | **Narrative**<br>(Briefly describe any key observations) |
| --- | --- | --- |
| **1) Are health workers at the PHC facility aware of the case definition of the pathogen in question?**<br>Randomly select a health worker to answer | Yes | HCWs have knowledge of case definitions, for measles and community members have high awareness of symptoms of measles |
| **2) Are health workers at the PHC facility aware of how and to whom to notify any further cases?**<br>Randomly select a health worker to answer | Yes | HCWs know the mean of notification e.g. phone call, SMS, WhatsApp eCBDS and they have contacts of their district IDSR focal persons |
| **3) Are health workers at the PHC facility aware of the IPC precautions needed for the pathogen in question?**<br>Randomly select a different health worker to answer | Yes | HCWs are aware of the IPC precautions needed for measles, but the supplies are not adequate |
| **4) Are health workers at the PHC facility aware of the clinical care/case management needed for the pathogen in question?**<br>Randomly select a health worker to answer | Yes | HCWs are aware of the clinical care/case management for measles due to frequent occurrence |
| **5) Does the PHC facility have a system for screening and triage to identify further cases?**<br>Visually confirm | Yes | HF has space for the activation of triage and screening area, thermometers, basic IPC system particularly for hand hygiene, relevant registers, and staffing. |
| **6) Does the PHC facility have a dedicated space for a holding area/isolation area for cases if required, or a process for temporary isolation?**<br>Visually confirm | Yes | HF has a dedicated space for isolation utilized as required. The HFs saw the majority of cases (49/52) and utilized their isolation area. |
| **7) Are health workers able to adapt the clinical setting to create more space for patients and/or move patients around (i.e., one-way flow of patients) the clinical setting safely during a surge?** | Yes | The HF's compound is fenced with an entry and exit point to enable one way flow of patients; the space is big enough to accommodate more patients. The HF itself has an entry and exit point |
| **8) Does the PHC facility have established mechanisms to engage with and rapidly disseminate information to their surrounding communities?**<br>Ask PHC Facility in-charge | Yes | HF disseminated information to their catchment communities through community healthcare workers (CHWs) and through HCWs follow up visits to the community |
| **9) Does the PHC facility have an up-to-date referral pathway for further cases?** | No | HF does not have a documented up-to-date referral pathway |
| **10) Does the PHC facility have adequate supplies to manage further cases?**<br>This should include a review of the PPE items needed, as well as drugs/consumables needed for case management | No | HF do not have adequate supplies to manage further cases |
| **Summary and Actions**<br>Review the responses to sections 1–3 to complete this summary section. Results from the overall assessment should be utilized to develop an action plan for improvement with the PHC facility staff. |  |  |
|  |  | **Yes/No** |
| Did the PHC facility meet the timeliness target for detection and notification?<br>Review answers to section 1 |  | Yes |
| Did the PHC facility complete all three of the immediate PHC facility actions correctly?<br>Review answers to section 2 |  | Yes |
| Is the PHC facility ready for a potential surge in cases?<br>Review answers to section 3 (all components should be present for yes) |  | No |
| **Action plan** |  |  |
| **Action** | **Responsible** | **Timeline** |
| Create community awareness on the need to visit HF early for treatment at onset of symptoms of diseases | In-charge |  |
| Activate community alert system | In-charge |  |
| Engage DHMTs to get adequate IPC supplies for potential case surge | In-charge |  |
| Develop an up-to-date referral pathway and list of referral HFs for surge in cases | In-charge |  |

**Annex 3:**
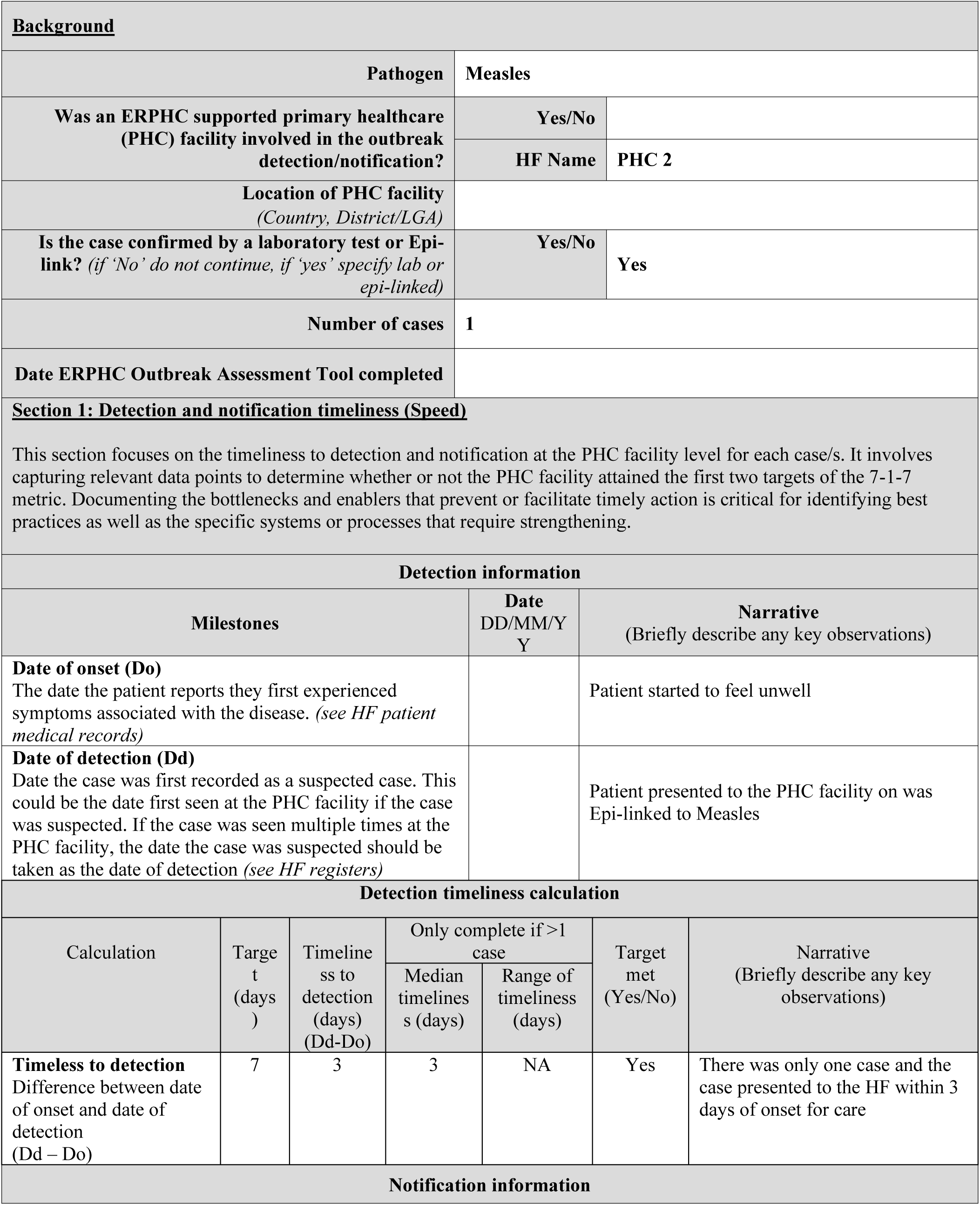
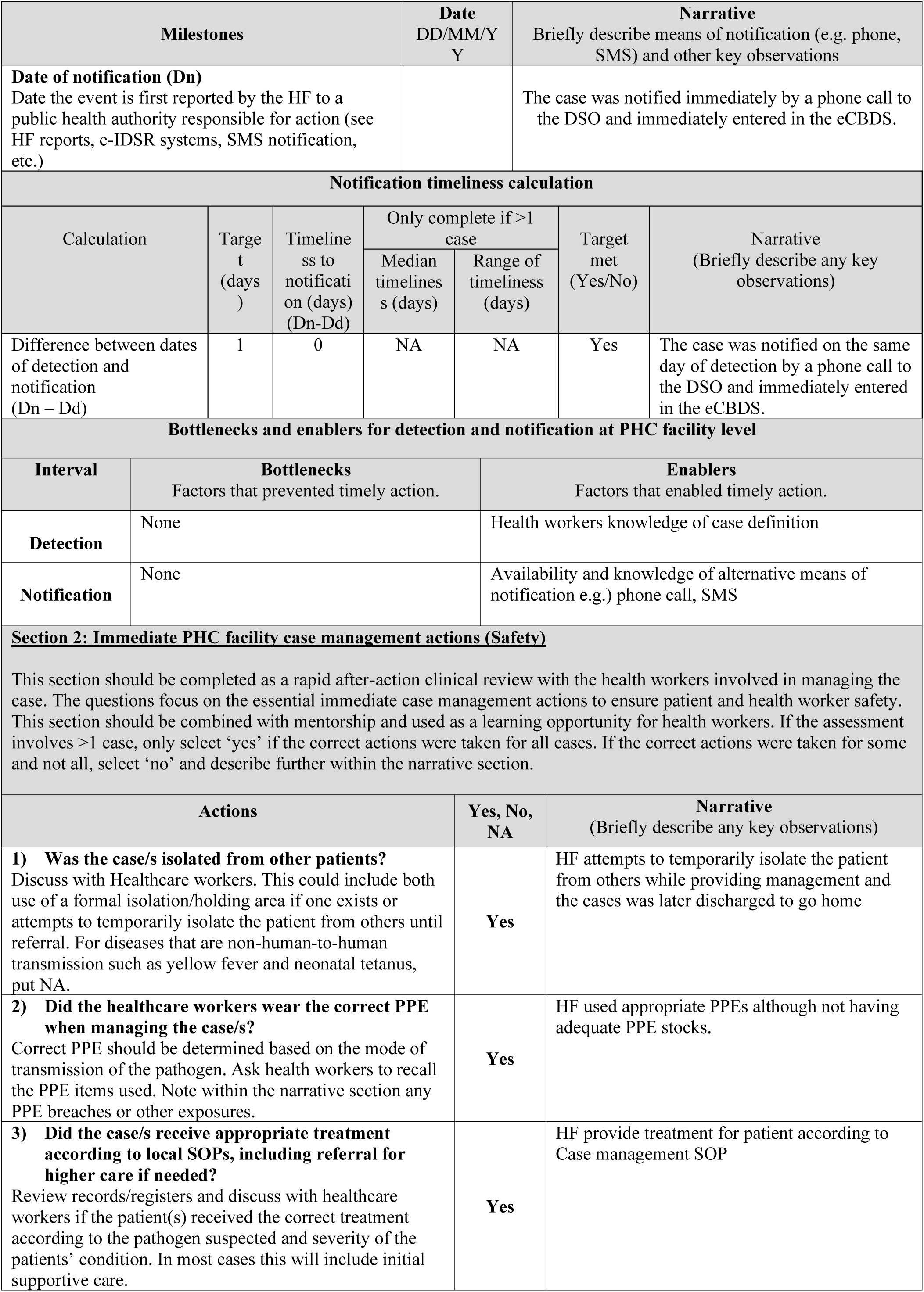
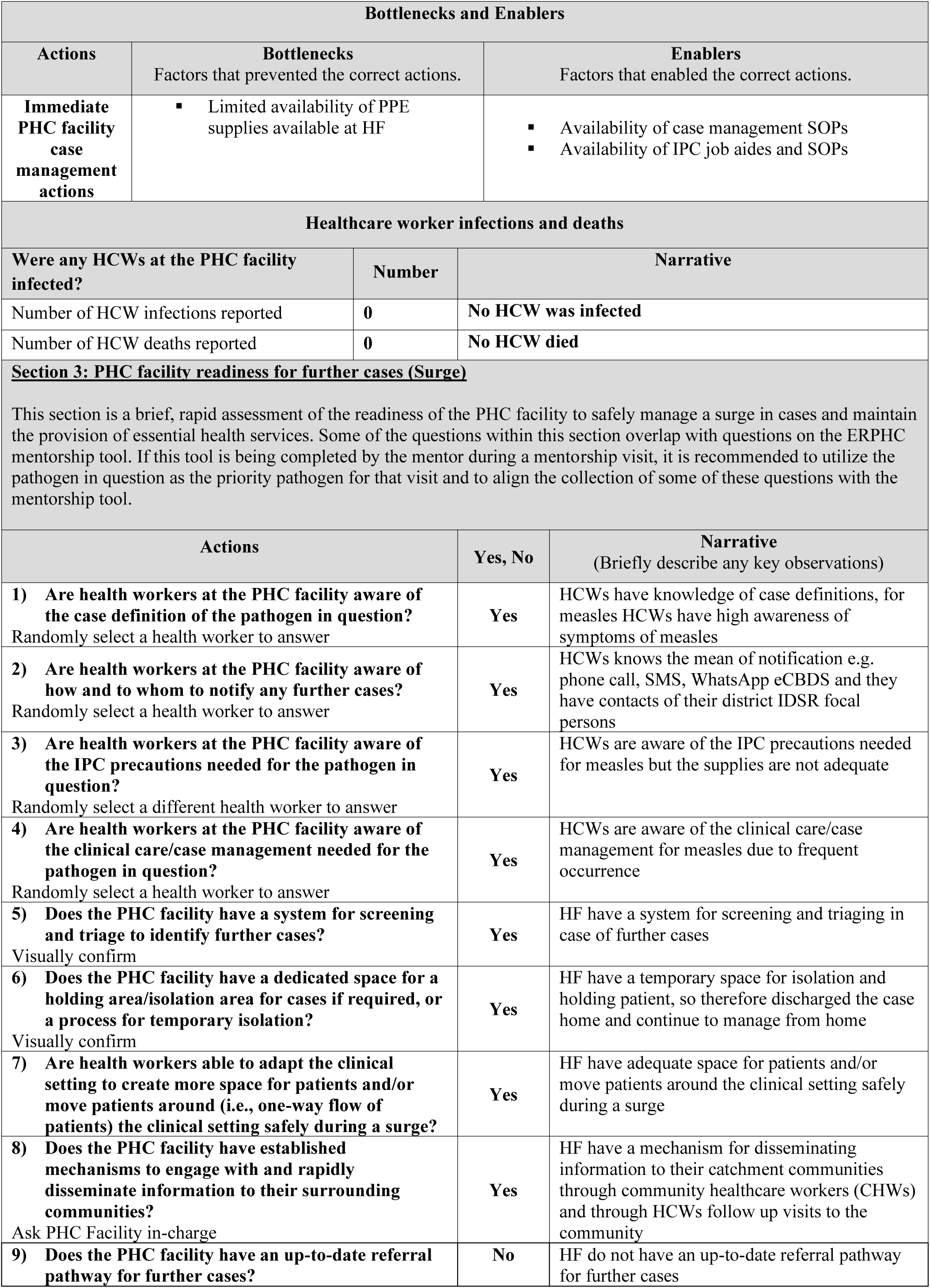
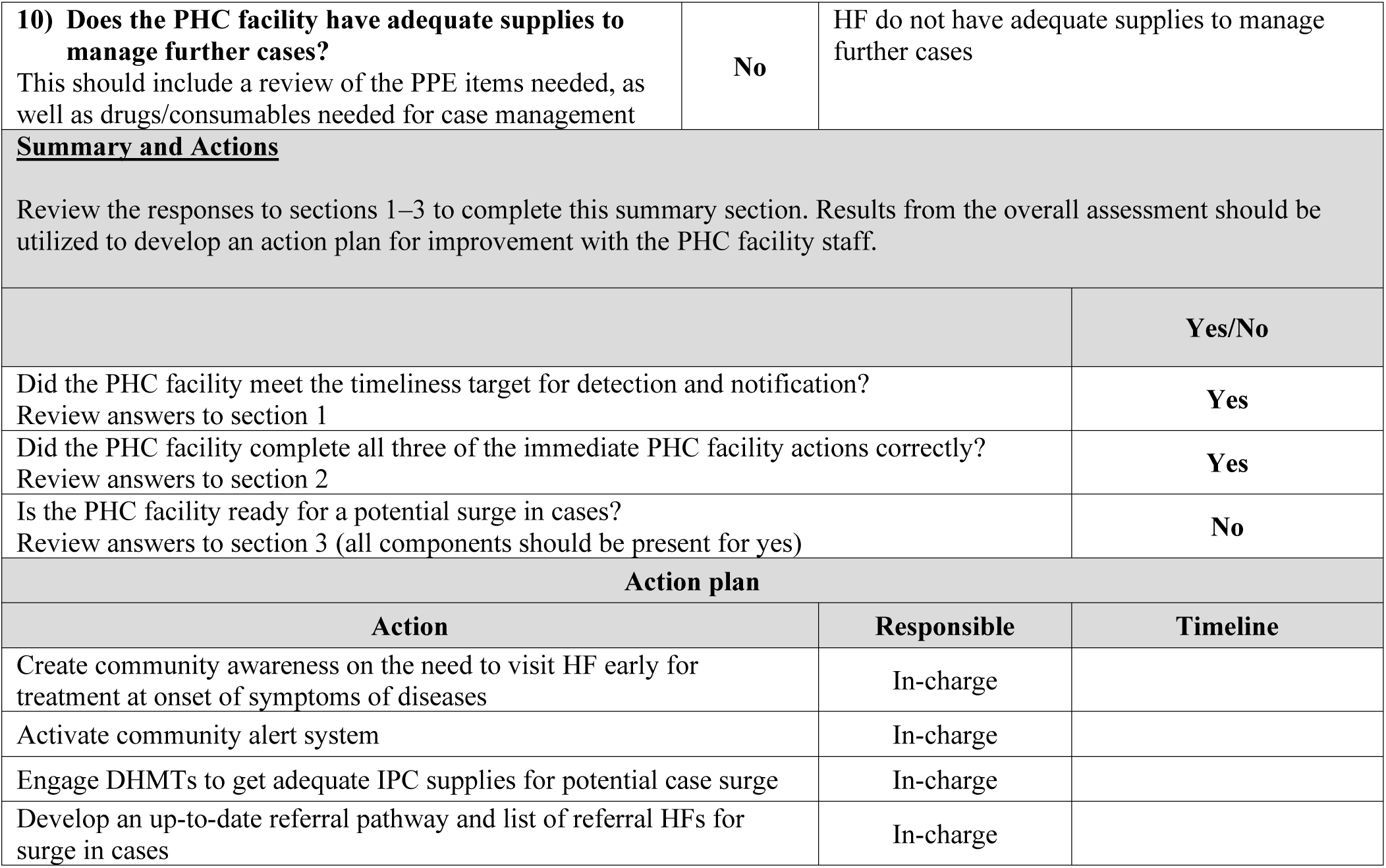
PHC 2 Measles Outbreak 7-1-7 Analysis Results.

**Annex 4:**
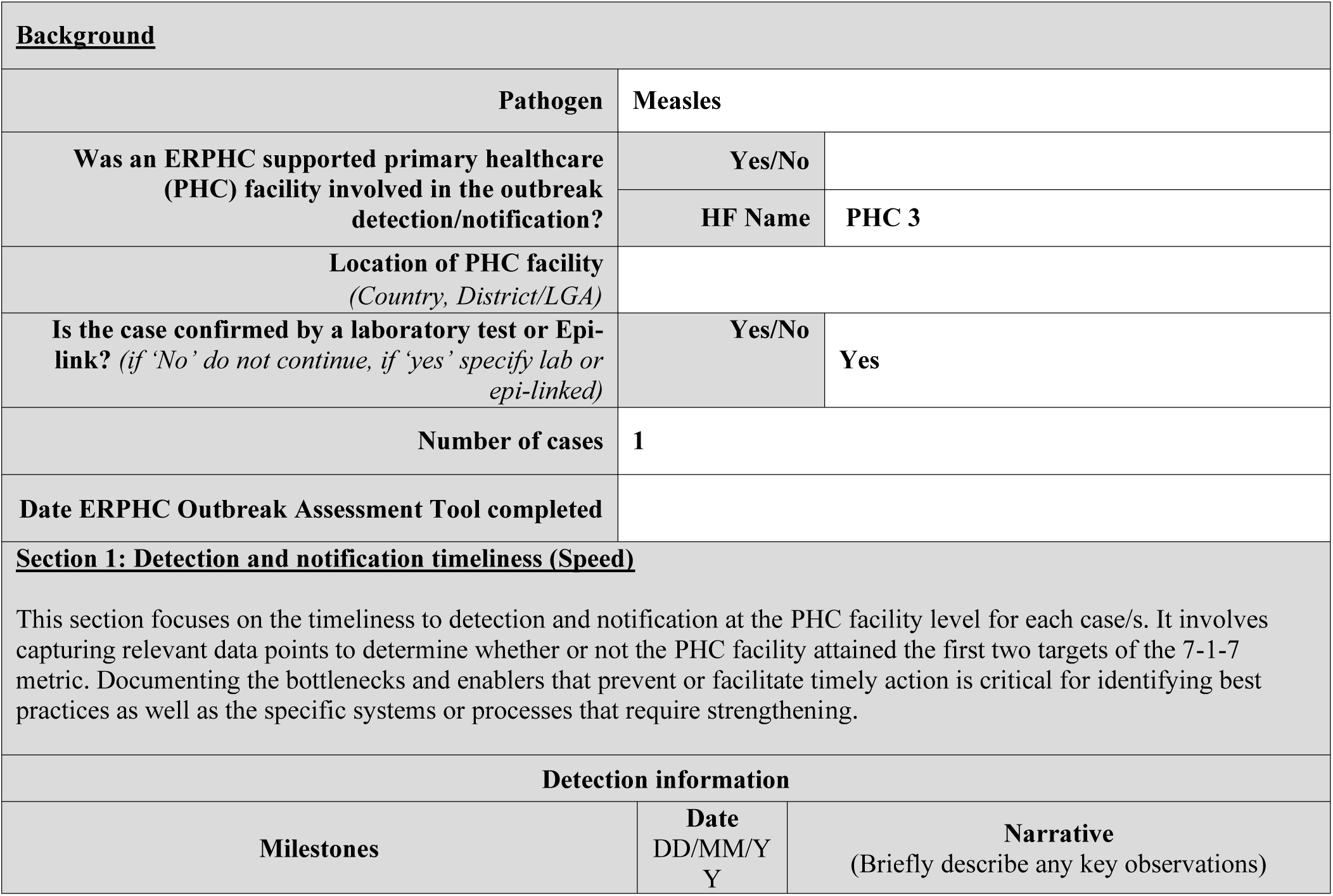
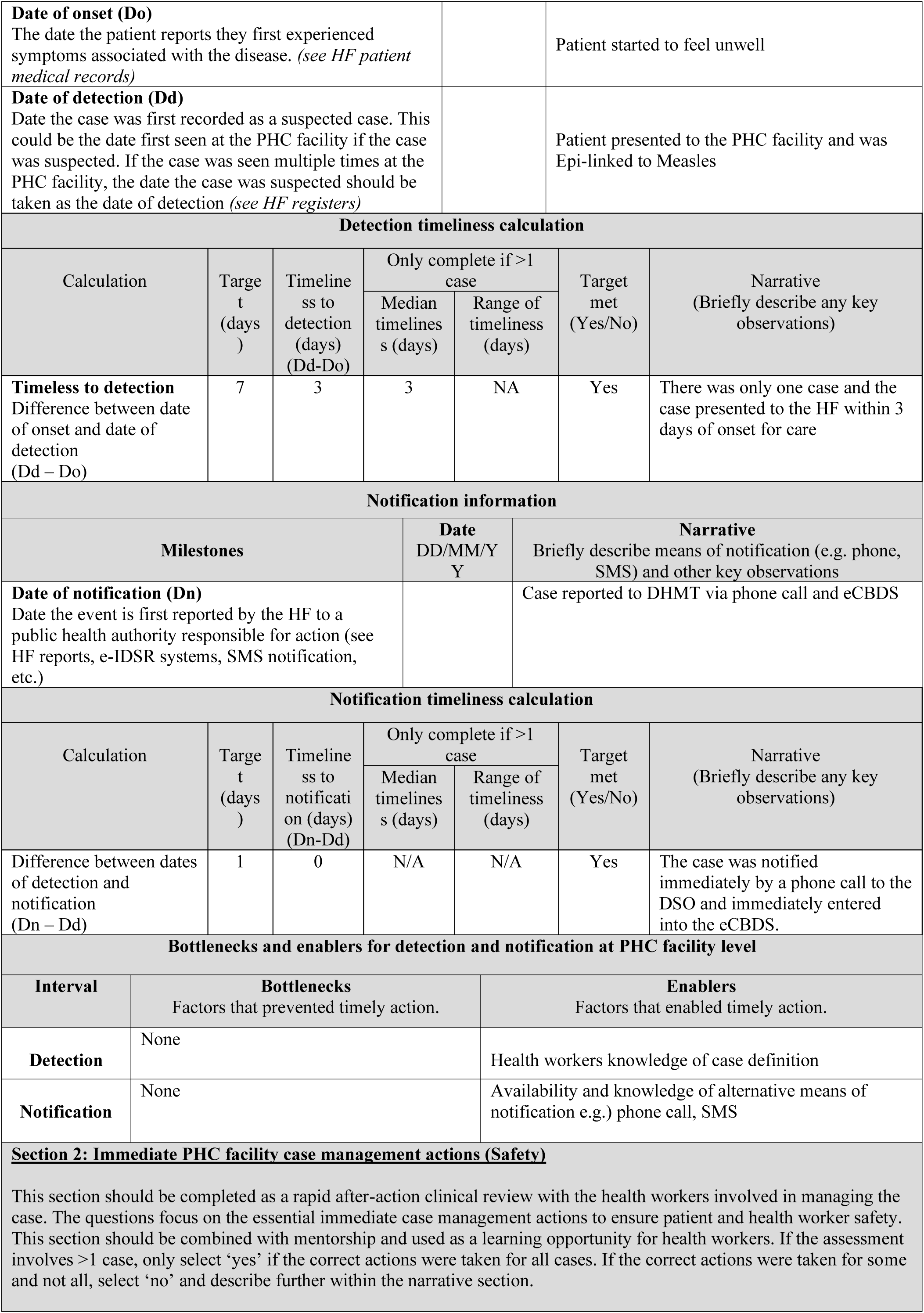
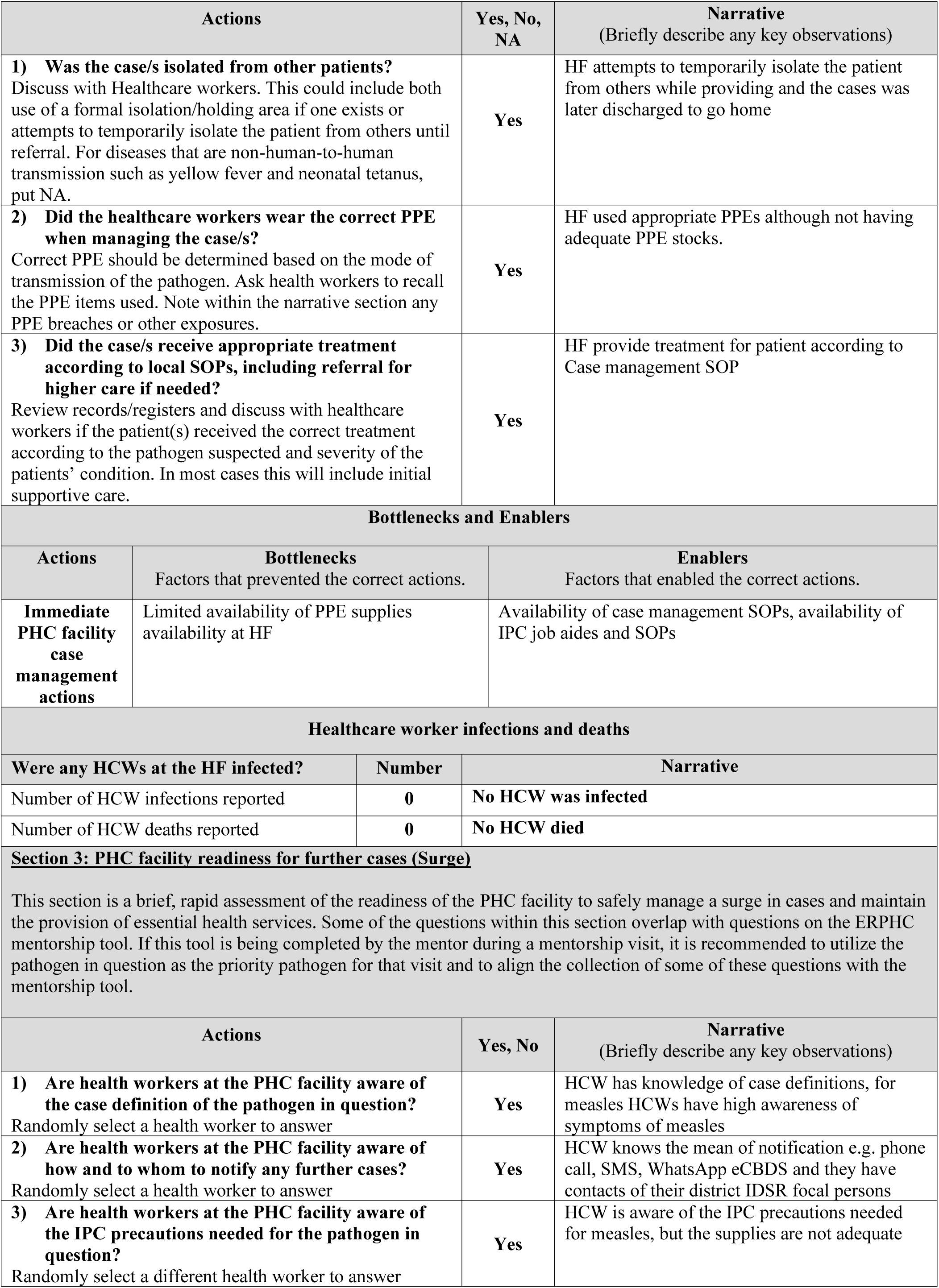
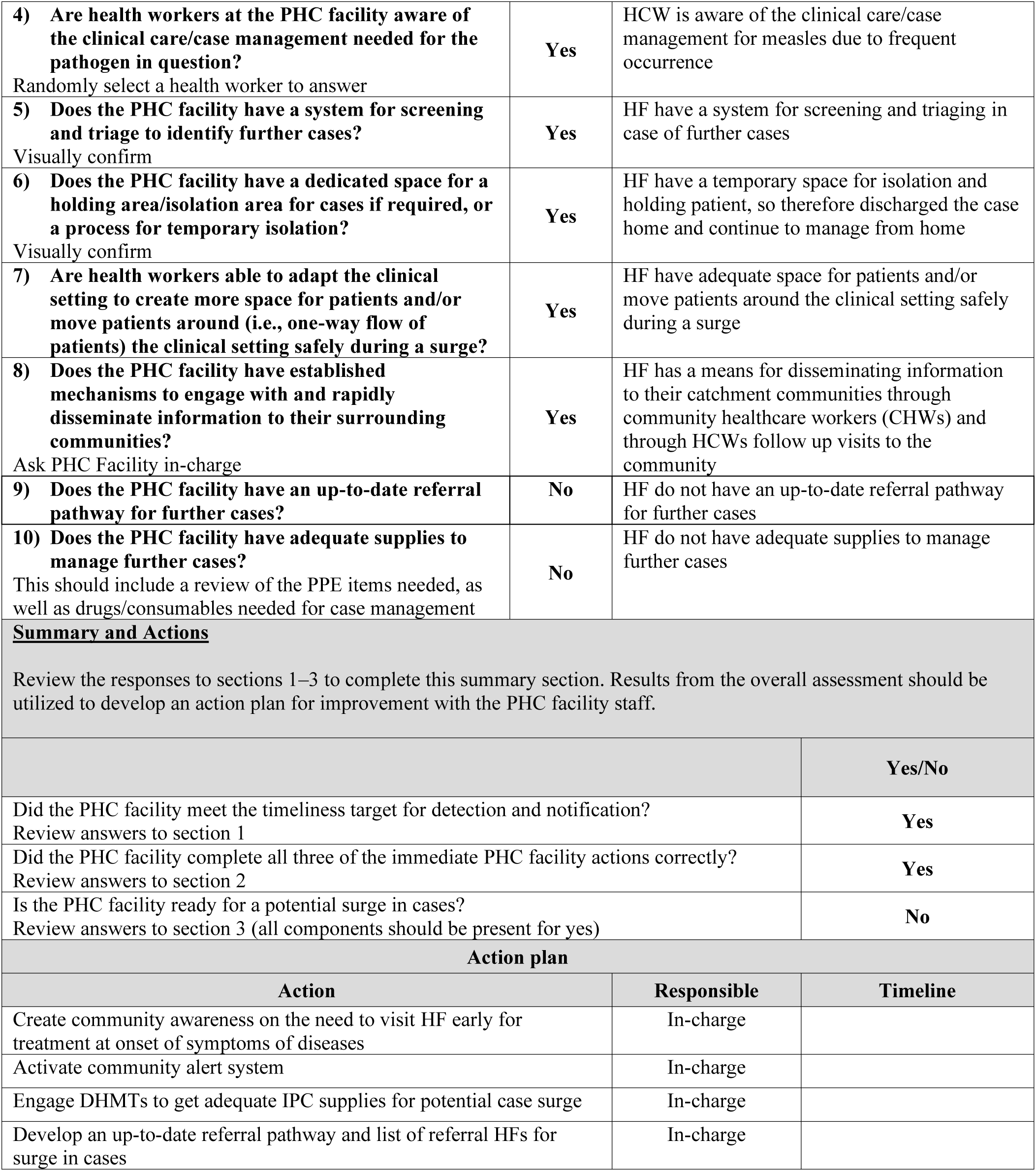
PHC 3Measles Outbreak 7-1-7 Analysis Results.

|  |  |  |
| --- | --- | --- |
| **<u>Background</u>** |  |  |
| **Pathogen** | Measles |  |
| **Was an ERPHC supported primary healthcare (PHC) facility involved in the outbreak detection/notification?** | **Yes/No** |  |
|  | **HF Name** | PHC 3 |
| **Location of PHC facility**<br>(Country, District/LGA) |  |  |
| **Is the case confirmed by a laboratory test or Epi-link?** (if 'No' do not continue, if 'yes' specify lab or epi-linked) | **Yes/No** | Yes |
| **Number of cases** | 1 |  |
| **Date ERPHC Outbreak Assessment Tool completed** |  |  |
| **<u>Section 1: Detection and notification timeliness (Speed)</u>** |  |  |
| This section focuses on the timeliness to detection and notification at the PHC facility level for each case/s. It involves capturing relevant data points to determine whether or not the PHC facility attained the first two targets of the 7-1-7 metric. Documenting the bottlenecks and enablers that prevent or facilitate timely action is critical for identifying best practices as well as the specific systems or processes that require strengthening. |  |  |
| **Detection information** |  |  |
| **Milestones** | **Date**<br>DD/MM/Y<br>Y | **Narrative**<br>(Briefly describe any key observations) |

| **Date of onset (Do)**<br>The date the patient reports they first experienced symptoms associated with the disease. (*see HF patient medical records*) |  |  |  |  | Patient started to feel unwell |  |
| --- | --- | --- | --- | --- | --- | --- |
| **Date of detection (Dd)**<br>Date the case was first recorded as a suspected case. This could be the date first seen at the PHC facility if the case was suspected. If the case was seen multiple times at the PHC facility, the date the case was suspected should be taken as the date of detection (*see HF registers*) |  |  |  |  | Patient presented to the PHC facility and was Epi-linked to Measles |  |
| **Detection timeliness calculation** |  |  |  |  |  |  |
| Calculation | Target (days) | Timeline ss to detection (days) (Dd-Do) | Only complete if >1 case |  | Target met (Yes/No) | Narrative (Briefly describe any key observations) |
|  |  |  | Median timelines (days) | Range of timeliness (days) |  |  |
| **Timeless to detection**<br>Difference between date of onset and date of detection (Dd – Do) | 7 | 3 | 3 | NA | Yes | There was only one case and the case presented to the HF within 3 days of onset for care |
| **Notification information** |  |  |  |  |  |  |
| **Milestones** |  |  | **Date**<br>DD/MM/YYYY |  | **Narrative**<br>Briefly describe means of notification (e.g. phone, SMS) and other key observations |  |
| **Date of notification (Dn)**<br>Date the event is first reported by the HF to a public health authority responsible for action (see HF reports, e-IDSR systems, SMS notification, etc.) |  |  |  |  | Case reported to DHMT via phone call and eCBDS |  |
| **Notification timeliness calculation** |  |  |  |  |  |  |
| Calculation | Target (days) | Timeline ss to notification (days) (Dn-Dd) | Only complete if >1 case |  | Target met (Yes/No) | Narrative (Briefly describe any key observations) |
|  |  |  | Median timelines (days) | Range of timeliness (days) |  |  |
| Difference between dates of detection and notification (Dn – Dd) | 1 | 0 | N/A | N/A | Yes | The case was notified immediately by a phone call to the DSO and immediately entered into the eCBDS. |
| **Bottlenecks and enablers for detection and notification at PHC facility level** |  |  |  |  |  |  |
| Interval | **Bottlenecks**<br>Factors that prevented timely action. |  |  | **Enablers**<br>Factors that enabled timely action. |  |  |
| **Detection** | None |  |  | Health workers knowledge of case definition |  |  |
| **Notification** | None |  |  | Availability and knowledge of alternative means of notification e.g.) phone call, SMS |  |  |
| **Section 2: Immediate PHC facility case management actions (Safety)**<br>This section should be completed as a rapid after-action clinical review with the health workers involved in managing the case. The questions focus on the essential immediate case management actions to ensure patient and health worker safety. This section should be combined with mentorship and used as a learning opportunity for health workers. If the assessment involves >1 case, only select 'yes' if the correct actions were taken for all cases. If the correct actions were taken for some and not all, select 'no' and describe further within the narrative section. |  |  |  |  |  |  |

| Actions |  | Yes, No, NA | Narrative<br>(Briefly describe any key observations) |
| --- | --- | --- | --- |
| **1) Was the case/s isolated from other patients?**<br>Discuss with Healthcare workers. This could include both use of a formal isolation/holding area if one exists or attempts to temporarily isolate the patient from others until referral. For diseases that are non-human-to-human transmission such as yellow fever and neonatal tetanus, put NA. |  | Yes | HF attempts to temporarily isolate the patient from others while providing and the cases was later discharged to go home |
| **2) Did the healthcare workers wear the correct PPE when managing the case/s?**<br>Correct PPE should be determined based on the mode of transmission of the pathogen. Ask health workers to recall the PPE items used. Note within the narrative section any PPE breaches or other exposures. |  | Yes | HF used appropriate PPEs although not having adequate PPE stocks. |
| **3) Did the case/s receive appropriate treatment according to local SOPs, including referral for higher care if needed?**<br>Review records/registers and discuss with healthcare workers if the patient(s) received the correct treatment according to the pathogen suspected and severity of the patients' condition. In most cases this will include initial supportive care. |  | Yes | HF provide treatment for patient according to Case management SOP |
| **Bottlenecks and Enablers** |  |  |  |
| Actions | Bottlenecks<br>Factors that prevented the correct actions. | Enablers<br>Factors that enabled the correct actions. |  |
| **Immediate PHC facility case management actions** | Limited availability of PPE supplies<br>availability at HF | Availability of case management SOPs, availability of IPC job aides and SOPs |  |
| **Healthcare worker infections and deaths** |  |  |  |
| Were any HCWs at the HF infected? | Number | Narrative |  |
| Number of HCW infections reported | 0 | No HCW was infected |  |
| Number of HCW deaths reported | 0 | No HCW died |  |
| **Section 3: PHC facility readiness for further cases (Surge)** |  |  |  |
| This section is a brief, rapid assessment of the readiness of the PHC facility to safely manage a surge in cases and maintain the provision of essential health services. Some of the questions within this section overlap with questions on the ERPHC mentorship tool. If this tool is being completed by the mentor during a mentorship visit, it is recommended to utilize the pathogen in question as the priority pathogen for that visit and to align the collection of some of these questions with the mentorship tool. |  |  |  |
| Actions | Yes, No | Narrative<br>(Briefly describe any key observations) |  |
| **1) Are health workers at the PHC facility aware of the case definition of the pathogen in question?**<br>Randomly select a health worker to answer | Yes | HCW has knowledge of case definitions, for measles HCWs have high awareness of symptoms of measles |  |
| **2) Are health workers at the PHC facility aware of how and to whom to notify any further cases?**<br>Randomly select a health worker to answer | Yes | HCW knows the mean of notification e.g. phone call, SMS, WhatsApp eCBDS and they have contacts of their district IDSR focal persons |  |
| **3) Are health workers at the PHC facility aware of the IPC precautions needed for the pathogen in question?**<br>Randomly select a different health worker to answer | Yes | HCW is aware of the IPC precautions needed for measles, but the supplies are not adequate |  |
| 4) **Are health workers at the PHC facility aware of the clinical care/case management needed for the pathogen in question?**<br>Randomly select a health worker to answer | Yes | HCW is aware of the clinical care/case management for measles due to frequent occurrence |
| 5) **Does the PHC facility have a system for screening and triage to identify further cases?**<br>Visually confirm | Yes | HF have a system for screening and triaging in case of further cases |
| 6) **Does the PHC facility have a dedicated space for a holding area/isolation area for cases if required, or a process for temporary isolation?**<br>Visually confirm | Yes | HF have a temporary space for isolation and holding patient, so therefore discharged the case home and continue to manage from home |
| 7) **Are health workers able to adapt the clinical setting to create more space for patients and/or move patients around (i.e., one-way flow of patients) the clinical setting safely during a surge?** | Yes | HF have adequate space for patients and/or move patients around the clinical setting safely during a surge |
| 8) **Does the PHC facility have established mechanisms to engage with and rapidly disseminate information to their surrounding communities?**<br>Ask PHC Facility in-charge | Yes | HF has a means for disseminating information to their catchment communities through community healthcare workers (CHWs) and through HCWs follow up visits to the community |
| 9) **Does the PHC facility have an up-to-date referral pathway for further cases?** | No | HF do not have an up-to-date referral pathway for further cases |
| 10) **Does the PHC facility have adequate supplies to manage further cases?**<br>This should include a review of the PPE items needed, as well as drugs/consumables needed for case management | No | HF do not have adequate supplies to manage further cases |
| **<u>Summary and Actions</u>**<br>Review the responses to sections 1–3 to complete this summary section. Results from the overall assessment should be utilized to develop an action plan for improvement with the PHC facility staff. |  |  |
|  |  | **Yes/No** |
| Did the PHC facility meet the timeliness target for detection and notification?<br>Review answers to section 1 |  | **Yes** |
| Did the PHC facility complete all three of the immediate PHC facility actions correctly?<br>Review answers to section 2 |  | **Yes** |
| Is the PHC facility ready for a potential surge in cases?<br>Review answers to section 3 (all components should be present for yes) |  | **No** |
| **Action plan** |  |  |
| **Action** | **Responsible** | **Timeline** |
| Create community awareness on the need to visit HF early for treatment at onset of symptoms of diseases | In-charge |  |
| Activate community alert system | In-charge |  |
| Engage DHMTs to get adequate IPC supplies for potential case surge | In-charge |  |
| Develop an up-to-date referral pathway and list of referral HFs for surge in cases | In-charge |  |

**Annex 5:** PHC 4 Measles Outbreak 7-1-7 Analysis Results.

|  |  |  |
| --- | --- | --- |
| **<u>Background</u>** |  |  |
| **Pathogen** | **Measles** |  |
| **Was an ERPHC supported primary healthcare (PHC) facility involved in the outbreak detection/notification?** | **Yes/No** |  |
|  | **HF Name** | **PHC 4** |

| **Location of PHC facility**<br>(Country, District/LGA) |  |  |  |  |  |  |
| --- | --- | --- | --- | --- | --- | --- |
| **Is the case confirmed by a laboratory test or Epi-link?** (if 'No' do not continue, if 'yes' specify lab or epi-linked) |  | **Yes/No** |  | **Yes** |  |  |
| **Number of cases** |  | **1** |  |  |  |  |
| **Date ERPHC Outbreak Assessment Tool completed** |  |  |  |  |  |  |
| **Section 1: Detection and notification timeliness (Speed)** |  |  |  |  |  |  |
| This section focuses on the timeliness to detection and notification at the PHC facility level for each case/s. It involves capturing relevant data points to determine whether or not the PHC facility attained the first two targets of the 7-1-7 metric. Documenting the bottlenecks and enablers that prevent or facilitate timely action is critical for identifying best practices as well as the specific systems or processes that require strengthening. |  |  |  |  |  |  |
| **Detection information** |  |  |  |  |  |  |
| **Milestones** |  | **Date**<br>DD/MM/Y<br>Y |  | **Narrative**<br>(Briefly describe any key observations) |  |  |
| **Date of onset (Do)**<br>The date the patient reports they first experienced symptoms associated with the disease. (see HF patient medical records) |  |  |  | Patient started to feel unwell |  |  |
| **Date of detection (Dd)**<br>Date the case was first recorded as a suspected case. This could be the date first seen at the PHC facility if the case was suspected. If the case was seen multiple times at the PHC facility, the date the case was suspected should be taken as the date of detection (see HF registers) |  |  |  | Patient presented to the PHC facility and was suspected as possible Measles |  |  |
| **Detection timeliness calculation** |  |  |  |  |  |  |
| Calculation | Target<br>(days) | Timeline<br>ss to<br>detection<br>(days)<br>(Dd-Do) | Only complete if >1<br>case |  | Target<br>met<br>(Yes/No) | Narrative<br>(Briefly describe any key<br>observations) |
|  |  |  | Median<br>timelines<br>(days) | Range of<br>timeliness<br>(days) |  |  |
| **Timeless to detection**<br>Difference between date of onset and date of detection<br>(Dd – Do) | 7 | 2 | 2 | NA | Yes | There was only one case and was presented to the HF within 2 days of onset for care |
| **Notification information** |  |  |  |  |  |  |
| **Milestones** |  | **Date**<br>DD/MM/Y<br>Y |  | **Narrative**<br>Briefly describe means of notification (e.g. phone, SMS) and other key observations |  |  |
| **Date of notification (Dn)**<br>Date the event is first reported by the HF to a public health authority responsible for action (see HF reports, e-IDSR systems, SMS notification, etc.) |  |  |  | The case was notified immediately by a phone call to the DSO and immediately entered in the eCBDS. |  |  |
| **Notification timeliness calculation** |  |  |  |  |  |  |
| Calculation | Target<br>(days) | Timeline<br>ss to<br>notification<br>(days)<br>(Dn-Dd) | Only complete if >1<br>case |  | Target<br>met<br>(Yes/No) | Narrative<br>(Briefly describe any key<br>observations) |
|  |  |  | Median<br>timelines<br>(days) | Range of<br>timeliness<br>(days) |  |  |
| Difference between dates of detection and notification (Dn – Dd) | 1 | 0 | NA | NA | Yes | The case was notified immediately by a phone call to the DSO and immediately entered in the eCBDS. |

Bottlenecks and enablers for detection and notification at PHC facility level
| Interval | Bottlenecks<br>Factors that prevented timely action. | Enablers<br>Factors that enabled timely action. |
| --- | --- | --- |
| Detection | None | Health workers knowledge of case definition |
| Notification | None | Availability and knowledge of alternative means of notification e.g.) phone call, SMS |

| Actions | Yes, No, NA | Narrative<br>(Briefly describe any key observations) |
| --- | --- | --- |
| **1) Was the case/s isolated from other patients?**<br>Discuss with Healthcare workers. This could include both use of a formal isolation/holding area if one exists or attempts to temporarily isolate the patient from others until referral. For diseases that are non-human-to-human transmission such as yellow fever and neonatal tetanus, put NA. | Yes | HF attempts to temporarily isolate the patient from others while providing and the cases was later discharged to go home |
| **2) Did the healthcare workers wear the correct PPE when managing the case/s?**<br>Correct PPE should be determined based on the mode of transmission of the pathogen. Ask health workers to recall the PPE items used. Note within the narrative section any PPE breaches or other exposures. | Yes | HF used appropriate PPEs although not having adequate PPE stocks. |
| **3) Did the case/s receive appropriate treatment according to local SOPs, including referral for higher care if needed?**<br>Review records/registers and discuss with healthcare workers if the patient(s) received the correct treatment according to the pathogen suspected and severity of the patients' condition. In most cases this will include initial supportive care. | Yes | HF provide treatment for patient according to Case management SOP |

Bottlenecks and Enablers
| Actions | Bottlenecks<br>Factors that prevented the correct actions. | Enablers<br>Factors that enabled the correct actions. |
| --- | --- | --- |
| **Immediate PHC facility case management actions** | Limited availability of PPE supplies<br>availability at HF | ○ Availability of case management SOPs<br>▪ Availability of IPC job aides and SOPs |

Healthcare worker infections and deaths
| Were any HCWs at the HF infected? | Number | Narrative |
| --- | --- | --- |
| Number of HCW infections reported | 0 | No HCW was infected |
| Number of HCW deaths reported | 0 | No HCW died |

| Actions | Yes, No | Narrative<br>(Briefly describe any key observations) |
| --- | --- | --- |
| 1) **Are health workers at the PHC facility aware of the case definition of the pathogen in question?**<br>Randomly select a health worker to answer | Yes | HCW have knowledge of case definitions, for measles |
| 2) **Are health workers at the PHC facility aware of how and to whom to notify any further cases?**<br>Randomly select a health worker to answer | Yes | HCW knows the mean of notification e.g. phone call, SMS, WhatsApp eCBDS and they have contacts of their district IDSR focal persons |
| 3) **Are health workers at the PHC facility aware of the IPC precautions needed for the pathogen in question?**<br>Randomly select a different health worker to answer | Yes | HCW is aware of the IPC precautions needed for measles, but the supplies are not adequate |
| 4) **Are health workers at the PHC facility aware of the clinical care/case management needed for the pathogen in question?**<br>Randomly select a health worker to answer | Yes | HCW are aware of the clinical care/case management for measles due to frequent occurrence |
| 5) **Does the PHC facility have a system for screening and triage to identify further cases?**<br>Visually confirm | Yes | HF have a system for screening and triaging in case of further cases |
| 6) **Does the PHC facility have a dedicated space for a holding area/isolation area for cases if required, or a process for temporary isolation?**<br>Visually confirm | Yes | HF have a temporary space for isolation and holding patient, so therefore discharged the case home and continue to manage from home |
| 7) **Are health workers able to adapt the clinical setting to create more space for patients and/or move patients around (i.e., one-way flow of patients) the clinical setting safely during a surge?** | Yes | HF have adequate space for patients and/or move patients around the clinical setting safely during a surge |
| 8) **Does the PHC facility have established mechanisms to engage with and rapidly disseminate information to their surrounding communities?**<br>Ask PHC Facility in-charge | Yes | HF disseminated information to their catchment communities through community healthcare workers (CHWs) and through HCWs follow up visits to the community |
| 9) **Does the PHC facility have an up-to-date referral pathway for further cases?** | No | HF do not have an up-to-date referral pathway for further cases |
| 10) **Does the PHC facility have adequate supplies to manage further cases?**<br>This should include a review of the PPE items needed, as well as drugs/consumables needed for case management | No | HF do not have adequate supplies to manage further cases |

|  | Yes/No |
| --- | --- |
| Did the PHC facility meet the timeliness target for detection and notification?<br>Review answers to section 1 | Yes |
| Did the PHC facility complete all three of the immediate PHC facility actions correctly?<br>Review answers to section 2 | Yes |
| Is the PHC facility ready for a potential surge in cases?<br>Review answers to section 3 (all components should be present for yes) | No |
| **Action plan** |  |

| Action | Responsible | Timeline |
| --- | --- | --- |
| Create community awareness on the need to visit HF early for treatment at onset of symptoms of diseases | In-charge |  |
| Activate community alert system | In-charge /IDSR<br>Focal |  |
| Engage DHMTs to get adequate IPC supplies for potential case surge | In-charge/ IPC |  |
| Develop an up-to-date referral pathway and list of referral HFs for surge in cases | In-charge |  |

## Acknowledgements

We thank the Ministry of Health and National Public Health Agency in Sierra Leone for their efforts in implementing this initiative. Our collaborative partner Resolve to Save Lives for co-implementation support.

## Contributors

OE and AEBC conceived the paper and served as the primary authors. SMS and SM contributed to the conceptualization and provided substantial editorial input. OE, AN, RM, MT, AY contributed data to the manuscript. OE analyzed the data. All authors reviewed and edited the final draft of the manuscript. All authors were involved in the design and implementation of the ERPHC pilot.

## Funding

This work was funded #Startsmall. Funding was provided via Resolve to Save Lives.

## Competing interests

Not applicable

## Ethics statements

This work was determined to be exempt human subjects research by the Columbia University Institutional Review Board and Resolve to Save Lives Research Committee. Approvals to disseminate findings or IRB review and approval was obtained as required in each country.

## Patient consent for publication

Not applicable

## Provenance and peer review

Not commissioned, external peer review

## Data Availability Statement

All data relevant to the study are included in the article or uploaded as online supplemental information.

## Notes

### Competing Interest Statement

The authors have declared no competing interest.

### Author Declarations

The Ethics committee/IRB at ICAP at Columbia University in New York City, NY, United States waived ethical approval for this work. The Ethics committee/IRB, named the Sierra Leone Ethics and Scientific Review Committee, in Freetown, Sierra Leone waived approval for this work.

